# Pathway-specific Polygenic Risk Scores (PRS) identify OSA-related pathways differentially moderating genetic susceptibility to CAD

**DOI:** 10.1101/2021.04.27.21255520

**Authors:** Matthew O Goodman, Brian E Cade, Neomi Shah, Tianyi Huang, Hassan S Dashti, Richa Saxena, Martin K Rutter, Peter Libby, Tamar Sofer, Susan Redline

**Affiliations:** Division of Sleep and Circadian Disorders, Brigham and Women’s Hospital and Harvard Medical School, Boston, MA; Division of Sleep Medicine, Harvard Medical School, Boston, MA; Program in Medical and Population Genetics, Broad Institute, Cambridge, MA, USA; Icahn School of Medicine at Mount Sinai, New York, NY; Channing Division of Network Medicine, Brigham and Women’s Hospital and Harvard Medical School, Boston, MA; Center for Genomic Medicine, Massachusetts General Hospital, Boston, MA, USA; Department of Anesthesia, Critical Care and Pain Medicine, Massachusetts General Hospital and Harvard Medical School, Boston, MA, USA; Division of Diabetes, Endocrinology and Gastroenterology, School of Medical Sciences, Faculty of Biology, Medicine and Health, University of Manchester, Manchester Academic Health Science Centre, Manchester, United Kingdom; Diabetes, Endocrinology and Metabolism Centre, Manchester University NHS Foundation Trust, Manchester Academic Health Science Centre, Manchester, United Kingdom; Division of Cardiovascular Medicine, Department of Medicine, Brigham and Women’s Hospital, Harvard Medical School, Boston, MA, USA.

## Abstract

**Background:** Obstructive sleep apnea (OSA) and its features, such as chronic intermittent hypoxia (IH), may differentially affect specific molecular pathways and processes in the pathogenesis of coronary artery disease (CAD) and influence the subsequent risk and severity of CAD events. In particular, competing adverse (e.g. inflammatory) and protective (e.g. increased coronary collateral blood flow) mechanisms may operate, but remain poorly understood. We hypothesize that common genetic variation in selected molecular pathways influences the likelihood of CAD events differently in individuals with and without OSA, in a pathway-dependent manner.

**Methods:** We selected a cross-sectional sample of 471,877 participants from the UK Biobank, among whom we ascertained 4,974 to have OSA, 25,988 to have CAD, and 711 to have both. We calculated pathway-specific polygenic risk scores (PS-PRS) for CAD, based on 6.6 million common variants evaluated in the CARDIoGRAMplusC4D genome-wide association study (GWAS), annotated to specific genes and pathways using functional genomics databases. Based on prior evidence of involvement with IH and CAD, we tested PS-PRS for the HIF-1, VEGF, NFκB and TNF signaling pathways.

**Results:** In a multivariable-adjusted logistic generalized additive model, elevated PS-PRSs for the KEGG VEGF pathway (39 genes) associated with protection for CAD in OSA (interaction odds ratio 0.86, *p* = 6E-04). By contrast, the genome-wide CAD PRS did not show evidence of statistical interaction with OSA.

**Conclusions:** We find evidence that pathway-specific genetic risk of CAD differs between individuals with and without OSA in a qualitatively pathway-dependent manner, consistent with the previously studied phenomena whereby features of OSA may have both positive and negative effects on CAD. These results provide evidence that gene-by-environment interaction influences CAD risk in certain pathways among people with OSA, an effect that is not well-captured by the genome-wide PRS. These results can be followed up to study how OSA interacts with genetic risk at the molecular level, and potentially to personalize OSA treatment and reduce CAD risk according to individual pathway-specific genetic risk profiles.

## INTRODUCTION

Obstructive sleep apnea (OSA) is a common disorder characterized by recurrent episodes of hypoxemia during sleep, sleep fragmentation, and activation of the sympathetic nervous system, potentially stimulating pro-inflammatory and pro-thrombotic pathways that drive atherosclerosis [1]. Epidemiological studies implicate OSA as a potentially modifiable risk factor for cardiovascular disease (CVD), stimulating research aimed at quantifying the impact of OSA and its treatment on both coronary artery disease (CAD) and cerebrovascular disease [2]. While OSA increases the incidence of cerebrovascular disease, weaker and less consistent associations support associations between OSA and incident CAD [3]. Moreover, large clinical trials of the effect of positive airway pressure (PAP) treatment of OSA on composite CVD outcomes have provided equivocal results, in some cases with point estimates in the direction of increased risk in the treatment group [4–7].

Potential explanations for disappointing results of PAP RCTs include treatment non-adherence, non-optimal trial participant selection (e.g. advanced CVD at baseline) and limited statistical power [8, 9]. Another possibility is that putative CVD benefits of OSA treatment may vary across individuals, influenced by factors related to characteristics of OSA-related stress not captured by the traditional metric of OSA severity (apnea hypopnea index, AHI), as well as by underlying host characteristics [10, 11], including genetic predisposition to CVD and genetic variation within known OSA pathways [12, 13]. Moreover, some features of OSA might even prove protective for CVD in some individuals, and therefore treatment of OSA with PAP could increase CVD risk among those individuals [14]. This hypothesis derives support from human and animal studies showing that exposure to intermittent hypoxemia - a cardinal feature of OSA - can promote collateral blood flow, reflective of well-known effects of ischemic preconditioning [15].

The need to develop a precision medicine-informed approach to CVD risk stratification in OSA agrees with data indicating that OSA-related CVD risk varies across subgroups defined by clusters of phenotypic traits [16]. A central question, and the one that we address here, is whether genetic variation in CAD risk interacts with OSA features to modulate CAD risk in a pathway-dependent manner. We conceptualize repeated intermittent hypoxia (IH) and reoxygenation as a modifying environment for preexisting genetic risk for CAD, locating our analysis within a gene-by-environment (GxE) interaction paradigm. Our pathway-level GxE approach provides a new scale on which to study the genetic architecture of CAD susceptibility and its modification by OSA.

We build on prior work that (1) identified genetic markers for CAD risk involved in atherosclerosis, small vessel disease, inflammatory pathways, and angiogenesis, with evidence from genome-wide association studies (GWAS), expression studies, animal experiments and clinical trials [17–19]; and (2) showed the influence of IH on a variety of pathways (in positive and negative directions) relevant to CAD, including redox, inflammatory and angiogenic pathways [20].

Our approach considers effect modification of genetic risk, aggregated to the pathway level, and investigates pathway-specific polygenic risk scores (PS-PRS) for CAD. PRSs have demonstrated success in identifying high risk populations, but may perform poorly due to overly simplistic dimension reduction when compared with non-linear methods to summarize risk across many alleles [21, 22]. Here we consider a novel PRS approach, where we group markers by pathways to create multiple PS-PRSs aligned logically with genetic architecture, to facilitate investigation of GxE interaction at the pathway level.

We hypothesized that common genetic variation, in specific molecular pathways for CAD that are linked to the pathophysiology of OSA, influences the likelihood of CAD events differently in individuals with and without OSA, in a pathway-dependent manner. Our work was motivated by the potential for differential effects of OSA on multiple processes that impact CAD, such as inflammation and atherosclerosis, thrombosis, collateral vessel formation (via angiogenesis), control of coronary artery blood flow, and plaque disruption. Of central interest is the idea that OSA could be a “double-edged sword” with both positive or negative effects on CAD. In particular, intermittent hypoxia – a physiological stress characteristic of OSA – may exert either positive effects through ischemic preconditioning or negative effects by boosting atherosclerotic processes including inflammation [14, 15]. We pre-selected four regulatory pathways of interest based on our hypotheses of the impacts of chronic IH on angiogenic and inflammatory processes in CAD, the HIF-1, VEGF, NFκB and TNF signaling pathways. We focused on these four representative pathways for their recognized roles in these processes, which have been previously shown to involve differential expression of constituent genes in OSA [23–32].

By investigating whether OSA enhances, does not change, or ameliorates genetic risk of developing CAD among individuals with specific risk allele profiles within these pathways, we aim to provide new mechanistic insight into the biological effects of OSA on CAD. In addition, we explore the ability to identify patients with OSA who may have either protection against or increased risk of CAD events from their OSA status according to their specific genetic profile. Ultimately, we propose that this approach provides a novel step on the path toward understanding the complex role of OSA in CAD and formulating a personalized approach to treatment.

## METHODS

### UK Biobank

The UK Biobank (UKBB) cohort consists of 502,620 participants, recruited between 2006 and 2010 from across the United Kingdom, and is described elsewhere in detail [33]. Participants were assessed at baseline using questionnaires that included components for ascertaining medical conditions, lifestyle, and demographic information. Additional interviews were conducted by trained medical staff to assess medical history, health status and medication intake. We used available self-reported doctor-diagnosed medical conditions and ICD10 codes derived from the UKBB hospital episode inpatient data set (retrieved 03/21/2019) to define medical conditions of interest. Participants were genotyped on two closely related genotyping arrays: UK BiLEVE: N = 49,939, and UK Biobank Axiom: 438,343. In total, 488,282 participants were genotyped. All genotypes were then imputed using reference sequence data as previously described [34]. The National Health Service National Research Ethics Service (ref. 11/NW/0382) gave approval for the UK Biobank study, with each participant providing written informed consent.

### OSA status assessment

We defined cases and controls as follows: Cases of OSA must have had self-reported snoring AND at least one of the following sleep apnea diagnoses:

1) an ICD10^1^ code G47.3 for sleep apnea (N = 6643) OR
2) a self-report of doctor diagnosed sleep apnea documented during an interview with a trained medical professional (UKBB field 20002, coding 1123: sleep apnoea) (N = 1757).

Controls were non-cases. Individuals who could not be confidently defined as either cases or controls were excluded. To reduce the possibility of including cases of central sleep apnea ascertained using sleep apnea diagnostic codes, we required snoring (a cardinal symptom of OSA), documented by self-report in the UKBB sleep questions interview. Among participants without OSA we excluded those who self-reported both snoring and daytime sleepiness, two common symptoms of OSA. A total of 471,877 participants (4,974 OSA cases) with complete covariate data were available after making these exclusions.

### CAD status assessment

Coronary artery disease (CAD) cases were assessed based on a composite definition, including UKBB participants with self-reported heart attack/myocardial infarction, phecode 411.2 or UKBB algorithmic ascertainment for myocardial infarction, including self-report, hospital admission, or death certificate.

### Covariate Assessment

Diagnostic information for medical conditions were obtained from the UKBB data sets for self-reported doctor-diagnosed non-cancer illness and hospital episode inpatient diagnostic codes. ICD10 codes were collapsed to medically interpretable groupings using the phecode system [35], using Phecode Map version 1.2 for WHO ICD10’s. Hypertension, type 1 (T1D) and type 2 diabetes (T2D), COPD, asthma, and pulmonary fibrosis were ascertained as composite phenotypes, as described in Supplementary Table S1. Additional covariates were self-reported race, genotype platform (BiLEVE vs Axiom) and UK Biobank-provided genetic PCs.

### Polygenic Risk Score Construction

We based our analysis of genetic CAD risk on a published PRS described by Khera *et al* [36], with single nucleotide variant (SNV) effects estimated for each of 6,630,150 SNVs, using the linkage disequilibrium (LD) aware PRS algorithm LDPred [37] and summary statistics from the CARDIoGRAMplusC4D Consortium CAD GWAS of 60,801 cases and 123,504 controls of mainly European ancestry [38].

Pathway specific polygenic risk scores (PS-PRSs) were calculated for each individual as the effect-weighted sum of the count of risk alleles at each locus across all pathway SNVs:

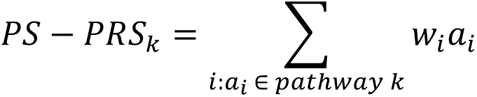

where *w_i_* is the LD-adjusted additive genetic effect and *a_i_* is the count (or imputed dosage) of the risk alleles at locus *i* of pathway *k*. PS-PRSs for select biological pathways were derived based on subsets of SNVs assigned to various pathways. Pathways are defined as sets of genes, and SNVs are assigned to genes via annotations derived from external reference data sets. We annotated SNVs in sequential rounds, prioritizing annotations with high specificity. SNVs assigned in prior rounds were set aside and were not annotated with less-specific data (Figure 1). We ordered the resources as follows: GenCode v34 exons [39], FANTOM5 [40], Promoter-Capture HiC from white blood cells [41], GTEx v7 eQTLs from artery, heart and whole blood tissues [42], GeneHancer v4.4 [43], and GenCode protein-coding regions (20kbp upstream to 10kbp downstream) and noncoding gene sequence regions. Further detail is given in the Methods Supplement. We also calculated gene-specific risk scores, GSRS*_j_*, i.e. PS-PRSs for a single gene, where the above sum is taken over loci *i*: *a_i_* ∈ *gene j* for a single gene of interest.

**Figure 1:**
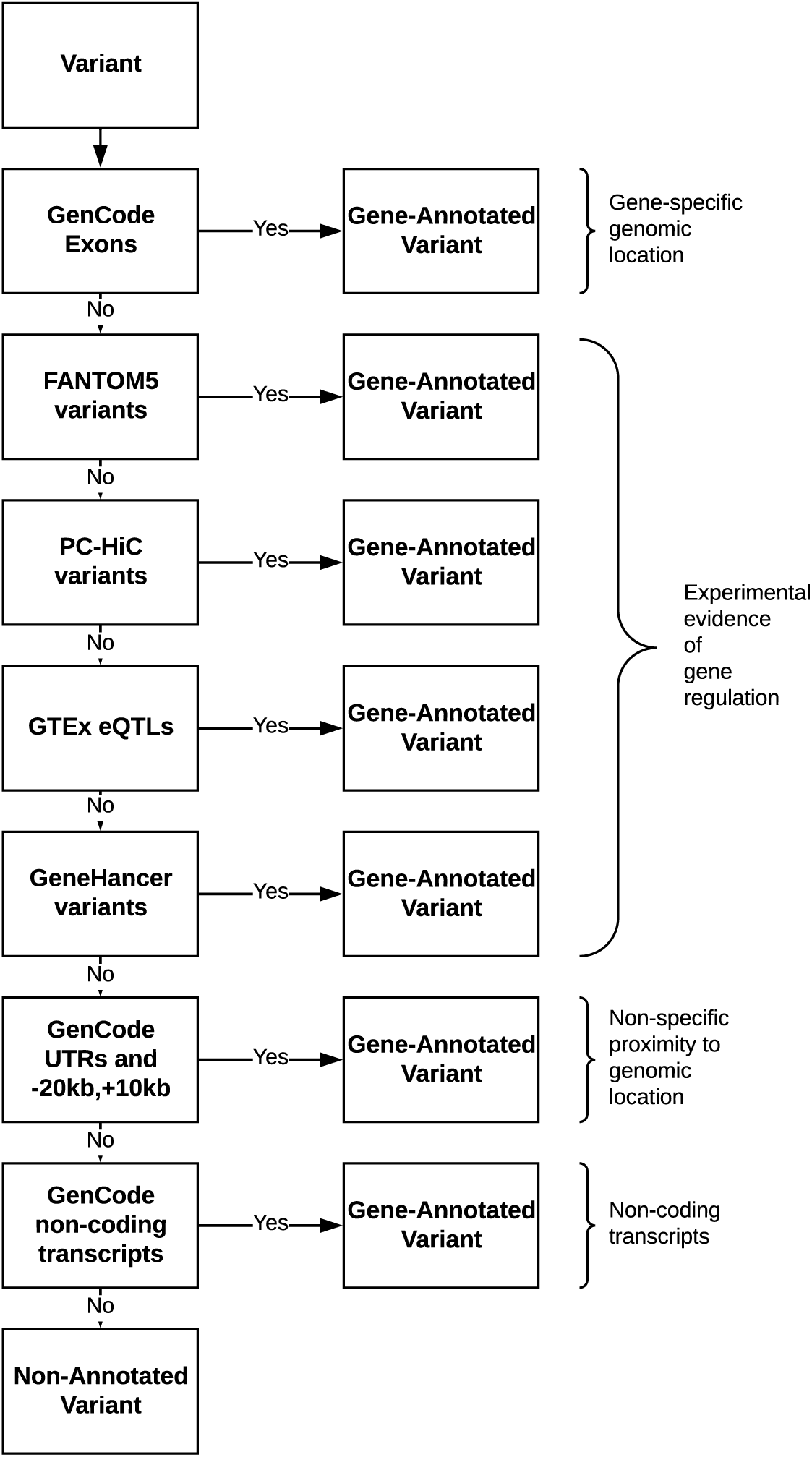
Flow diagram showing the gene annotation process.

### Defining pathways of interest

Based on our *a priori* hypotheses on the role hypoxemia plays in CAD, inducing angiogenesis and inflammation, we selected four pathways from the literature, for which the nominal genes, namely *HIF1A*, *VEGFA*, *NFKB1/NFKB2* and *TNF*, have been shown to be differentially expressed in OSA. For each, we created gene-pathways using two approaches. First, we selected the core-gene pathways, defined as the nominal gene(s) and their receptors (for HIF1, core genes selected were: *HIF1A* and *ARNT*, for VEGF: *VEGFA, FLT,* and *KDR*, for NFκB: *NFKBIA*, *NFKB1*, *RELA*, *NFKB2*, *RELB*, and for TNF: *TNF*, *TNFRSF1A*, and *TNFRSF1B*). Second, we selected more comprehensive gene pathways based the Kyoto Encyclopedia of Genes and Genomes (KEGG) database: HIF-1 signaling (KEGG pathway code: hsa04066), VEGF signaling (hsa04370), NFκB signaling (hsa04064), and TNF signaling (hsa04668). We also included, as comparators, the full PRS on 6.6M SNVs, as well as a curated CAD/OSA pathway consisting of 225 genes involved in both OSA and CAD based on literature review. Because KEGG pathway boundaries are somewhat arbitrary and include varying levels of completeness in upstream and downstream gene modules, we also organized each KEGG pathway into sub-modules. For example, the KEGG VEGF pathway was organized into several partially overlapping downstream submodules according to the KEGG pathway diagram, including: endothelial proliferation (15 genes), endothelial migration (9 genes) and endothelial survival (12 genes). For complete pathway and module definitions see Supplementary Table S2.

### Statistical Analysis

We studied the interaction of OSA case status with pathway-specific risk for CAD in several steps. 1) We attempted to annotate each marker identified by Khera *et al* to one or more associated genes (Figure 1). SNV markers that were not annotated were discarded, resulting in a QC-filtered, gene-annotated, genome-wide set of markers. 2) We selected pathways of interest and defined pathway-based SNV-sets implied by the SNV-gene and gene-pathway assignments. 3) We computed pathways-specific genetic risk scores (PS-PRS), as a sum of allele counts weighted by per-allele effect size and direction across the pathway SNV-set. 4) We tested the associations of PS-PRS and OSA status, and their GxE interaction, on the composite CAD outcome, in a logistic generalized additive model, controlling for the mutual interaction of age and BMI by sex (via tensor-product splines) as well as smoking and its interaction with sex [44], self-reported white race, the first 5 genetic PCs, genotype platform (BiLEVE), asthma and chronic obstructive pulmonary disease (COPD). As two of the most common pulmonary diseases, COPD and asthma were controlled for as both are associated with both OSA and CAD, and can potentially affect gas exchange and exacerbate hypoxia. The ten pathways of interest described were allocated Type-1 error in the primary analysis, with Bonferroni-corrected significance level of 5E-3. In a secondary analysis, to further explore whether module and gene-level PRS showed consistent associations with CAD, as well as to suggestively localize effects estimated in the primary analysis, we looked at pathway sub-modules and individual genes. For each core-gene module we modeled GxE interaction for the gene-specific risk scores (GSRS) of the constituent genes, and for each KEGG pathway, we modeled sub-module PS-PGRSs and GSRSs and reported those with both nominally significant risk-amplifying main effects and nominally significant GxE interactions. We further performed the following sensitivity analyses: including comorbidity covariates potentially mediating the OSA-CAD relationship (hypertension, Type 1 and Type 2 diabetes, along with their sex-interactions [44], and pulmonary fibrosis); excluding individuals with heart failure or stroke; sex-stratified analysis; and an analysis limited to self-reported white Europeans (the predominant ancestry in the CAD GWAS). Further detail is provided in the supplement.

## RESULTS

### Clinical characteristics

Of the 476,851 participants in our analytic sample, 4,974 (1.1%) were classified to have OSA and 26,699 (5.6%) to have CAD. The median (IQR) age was 58 (50, 63) years with a similar age distribution by OSA status (Table 1).

**Table 1.** Sample characteristics. Outcome, covariate and polygenic risk score distributions by OSA status. OSA: Obstructive sleep apnea, CAD: coronary artery disease, BMI: body mass index, PC: principal component, COPD: chronic obstructive pulmonary disease, KEGG: Kyoto encyclopedia of genes and genomes, hsa: Homo sapiens pathway reference number, HIF1: hypoxia inducible factor 1, VEGF: vascular endothelial growth factor, NFκB: nuclear factor kappa-beta, TNF: tumor necrosis factor, PRS: polygenic risk score, PS-PRS: pathway-specific polygenic risk score.

|  | OSA control |  | OSA case |  |
| --- | --- | --- | --- | --- |
| N | 471,877 |  | 4,974 |  |
| **Outcome:** |  |  |  |  |
| CAD, N (%) | 25,988 | (5.50) | 711 | (14.30) |
| **Covariates:** |  |  |  |  |
| BMI (median [IQR]) | 26.67 | (24.09, 29.79) | 31.61 | (28.11, 36.39) |
| Age (median [IQR]) | 58 | (50.00, 63.00) | 58 | (51.00, 63.00) |
| Male (%) | 213,700 | (45.30) | 3,622 | (72.80) |
| Smoking (%) |  |  |  |  |
| current | 36,568 | (7.70) | 519 | (10.40) |
| never | 259,572 | (55.00) | 2,162 | (43.50) |
| occasionally | 12,484 | (2.60) | 183 | (3.70) |
| previous | 163,253 | (34.60) | 2,110 | (42.40) |
| Self-reported white (%) | 397,269 | (84.20) | 4,054 | (81.50) |
| Genetic PC1 (mean (SD)) | -1.53 | (53.33) | 1.95 | (60.96) |
| Genetic PC2 (mean (SD)) | 0.5 | (27.38) | 0.21 | (27.29) |
| Genetic PC3 (mean (SD)) | -0.17 | (14.61) | 0.82 | (15.30) |
| Genetic PC4 (mean (SD)) | 0.13 | (10.32) | -0.2 | (12.07) |
| Genetic PC5 (mean (SD)) | 0.05 | (7.57) | 0.28 | (7.42) |
| BiLEVE genotype platform (%) | 48,528 | (10.30) | 619 | (12.40) |
| COPD (%) | 18,888 | (4.00) | 608 | (12.20) |
| Asthma (%) | 62,630 | (13.30) | 1,126 | (22.60) |
| **Standardized PRS and PS-PRS Exposures:** |  |  |  |  |
| KEGG hsa04066 HIF1 (mean (SD)) | -0.0002 | 1.000 | -0.011 | 0.985 |
| KEGG hsa04370 VEGF (mean (SD)) | -0.0014 | 1.000 | 0.029 | 0.982 |
| KEGG hsa04064 NFκB (mean (SD)) | 0.0002 | 1.000 | 0.003 | 1.005 |
| KEGG hsa04668 TNF (mean (SD)) | 0.0002 | 1.000 | 0.015 | 1.017 |
| HIF1 core genes (mean (SD)) | 0.0018 | 1.000 | -0.016 | 0.999 |
| VEGF core genes (mean (SD)) | 0.0003 | 1.000 | -0.010 | 0.991 |
| NFκB core genes (mean (SD)) | 0.0011 | 1.000 | -0.015 | 1.006 |
| TNF core genes (mean (SD)) | -0.0009 | 1.000 | 0.001 | 0.992 |
| CAD PRS (mean (SD)) | 0.0000 | 0.998 | 0.034 | 1.015 |
| OSA genes (mean (SD)) | -0.0016 | 0.999 | -0.005 | 0.997 |

As expected, OSA cases tended to have a higher prevalence of several CAD risk factors and comorbidities. Individuals classified with OSA were predominantly male, had a higher mean BMI, and included more ever-smokers. Pulmonary diseases, including COPD and asthma also tended to have a higher prevalence among OSA cases (Table 1). There was a higher percentage of OSA cases genotyped on the BiLEVE platform, as opposed to the UKBB Axiom platform, consistent with the fact that the BiLEVE pilot study was enriched for pulmonary disease.

### Characteristics of the PS-PRSs

Our annotation procedure resulted in the assignment of 70.3% of the 6.6M markers to one or more genes (average: 1.1 genes assigned per annotated variant). Pathway genes are shown in Supplementary Table S2. Gene-annotated CAD PRS data is available on request. After standardization on the cohort as a whole, participants with or without OSA had similar PS-PRS (Table 1).

The NFκB signaling pathway (KEGG hsa04064: 100 genes) and TNF signaling pathway (KEGG hsa04668: 110 genes) share 29 genes, which induced a correlation of 0.39 in the pathway-specific risk scores (Figure 2). The curated 225-gene OSA/CAD pathway shares 22 genes with the KEGG HIF1 pathway, driving the observed correlation of 0.54. Similarly, because VEGF core-genes *VEGFA* and *FLT1* and HIF1 core-genes *HIF1A* and *ARNT* reside within the HIF1 KEGG pathway, they induce correlations of 0.44 and 0.32 respectively (Figure S3). Additional pairwise correlations for pathways, genes, and modules shown in Figures 2 and S3. Note that as pathways largely come from disjoint regions of the genome and, due to genetic recombination, allele counts from distal variants with low LD (e.g. in separate genes) will be nearly uncorrelated statistically. Hence the low observed correlation between participants’ genetic risk scores in the majority of pathway pairs is expected due to statistical independence of the non-overlapping variants, and conversely, observed correlations are driven by the PS-PRS components attributable to the overlapping genes.

**Figure 2:**
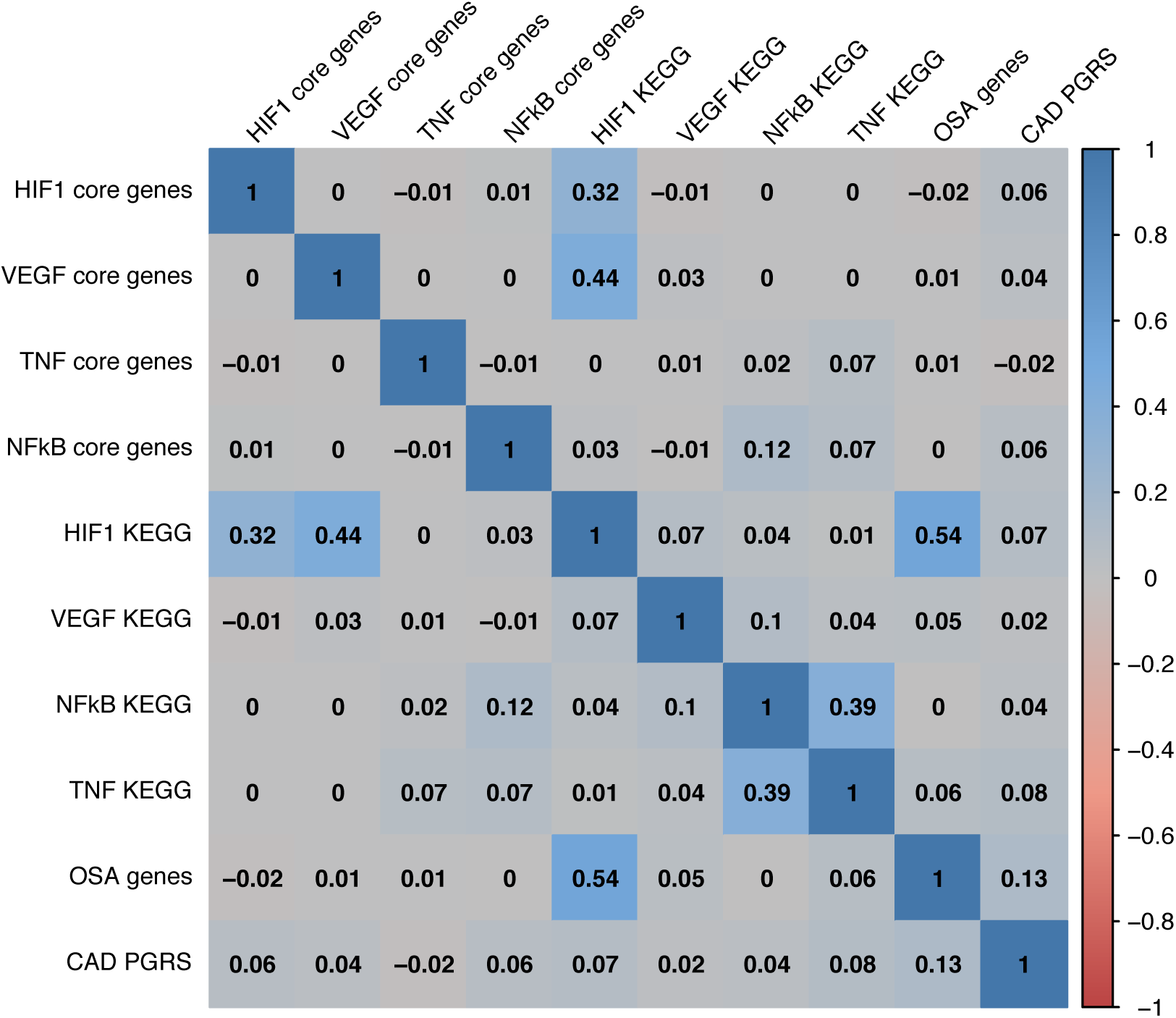
Pairwise correlations among selected pathway-specific polygenic risk scores. Pearson correlations between participants’ primary analysis risk scores.

### Estimated main effects of OSA, PRS and PS-PRSs

We found that OSA associated with higher CAD risk, with an approximate odds ratio (OR) of 1.5, and 95% confidence interval (CI) of (1.4, 1.6) across all models shown in Table 2. We also confirmed that the full genome-wide CAD PRS associated strongly with increased CAD risk in our model. For each standard deviation increase in the CAD PRS, the odds of CAD (in both OSA and OSA controls) increased by a factor of 1.70 (CI: 1.67, 1.72). Additionally, the PS-PRSs for each KEGG signaling pathway associated strongly with CAD, but had notably smaller estimated odds ratios (range 1.040 to 1.062). Core gene modules had still smaller odds ratios (range 1.019 to 1.032) (Table 2).

**Table 2:** Primary analysis of selected OSA pathways: Covariate-adjusted odds of CAD per s.d. of standardized PS-PRSs with effect modification by OSA status. In Table 2, each row depicts a separate PS-PRS model showing the OSA and PS-PRS main effects and the PS-PRS x OSA interaction effect in a model of the form logit(CAD) = OSA + PS-PRS + PS-PRSxOSA + covariates. Three groupings representing different types of pathways are included: A) core-gene modules B) KEGG-defined signaling pathways, and C) aggregate pathways. These ten pathways are allocated Type-1 error in the primary analysis. Nominally significant (*) GxE results do not pass Bonferroni-corrected significance (**) level 5E-3. Covariates adjusted in a logistic generalized additive model: mutual interaction of age and BMI by sex (via tensor-product splines) as well as smoking and its interaction with sex, self-reported white race, the first 5 genetic PCs, genotype platform (BiLEVE), asthma and chronic obstructive pulmonary disease. OR: odds ratio; CI: confidence interval; OSA: Obstructive sleep apnea, CAD: coronary artery disease, KEGG: Kyoto encyclopedia of genes and genomes, HIF1: hypoxia inducible factor 1, VEGF: vascular endothelial growth factor, NFκB: nuclear factor kappa-beta, TNF: tumor necrosis factor, PRS: polygenic risk score, PS-PRS: pathway-specific polygenic risk score.

| PS-PRS | OSA effect |  | PS-PRS effect in OSA Controls |  |  | PS-PRS effect in OSA Cases |  |  | OSA x PS-PRS interaction effect |  |
| --- | --- | --- | --- | --- | --- | --- | --- | --- | --- | --- |
|  | OR | CI | OR | CI | p-value | OR | CI | p-value | OR | p-value |
| **A) Selected pathways: core-gene modules** |  |  |  |  |  |  |  |  |  |  |
| HIF1 core genes | 1.50 | (1.38, 1.64) | 1.022 | (1.009, 1.035) | 1.06E-03 | 1.114 | (1.025, 1.212) | 1.10E-02 | 1.09 | 4.46E-02* |
| VEGF core genes | 1.51 | (1.38, 1.64) | 1.032 | (1.019, 1.045) | 1.66E-06 | 0.952 | (0.874, 1.037) | 2.58E-01 | 0.92 | 6.70E-02 |
| NFκB core genes | 1.51 | (1.38, 1.64) | 1.031 | (1.018, 1.045) | 4.45E-06 | 0.981 | (0.903, 1.066) | 6.54E-01 | 0.95 | 2.45E-01 |
| TNF core genes | 1.51 | (1.38, 1.64) | 1.019 | (1.006, 1.033) | 4.44E-03 | 1.013 | (0.931, 1.102) | 7.59E-01 | 0.99 | 8.94E-01 |
| **B) Selected pathways: KEGG signaling pathways** |  |  |  |  |  |  |  |  |  |  |
| HIF1 KEGG | 1.51 | (1.38, 1.64) | 1.062 | (1.048, 1.076) | 6.81E-20 | 1.089 | (1.001, 1.185) | 4.84E-02 | 1.03 | 5.66E-01 |
| VEGF KEGG | 1.51 | (1.38, 1.64) | 1.040 | (1.027, 1.054) | 4.30E-09 | 0.894 | (0.821, 0.974) | 1.02E-02 | 0.86 | 5.98E-04** |
| NFκB KEGG | 1.51 | (1.38, 1.64) | 1.058 | (1.044, 1.072) | 3.26E-17 | 0.998 | (0.919, 1.085) | 9.71E-01 | 0.94 | 1.74E-01 |
| TNF KEGG | 1.51 | (1.38, 1.64) | 1.052 | (1.038, 1.065) | 1.65E-14 | 0.994 | (0.916, 1.079) | 8.86E-01 | 0.95 | 1.85E-01 |
| **C) Selected aggregated pathway comparators** |  |  |  |  |  |  |  |  |  |  |
| CAD PRS | 1.52 | (1.39, 1.67) | 1.695 | (1.672, 1.719) | <1.0E-99 | 1.664 | (1.525, 1.816) | 3.30E-30 | 0.98 | 6.80E-01 |
| OSA genes | 1.50 | (1.37, 1.63) | 1.102 | (1.087, 1.116) | 1.17E-46 | 1.222 | (1.122, 1.330) | 3.81E-06 | 1.11 | 1.83E-02* |

### Estimated interaction effects between OSA, and PS-PRSs

In our primary analysis, we find evidence of effect-measure modification of genetic risk by OSA status. Specifically, at Bonferroni-corrected significance-level 5E-3, we see significant qualitative effect modification in the VEGF KEGG pathway (interaction odds ratio 0.86, *p* = 6E-04), leading to differential genetic effects among participants with and without OSA (Table 2). Concretely, among OSA cases, we estimated the odds of CAD decreased, by a factor of 0.89 per standard deviation increase in the VEGF KEGG PS-PRS, contrasted with an increase of 1.04 among OSA controls. We find a qualitatively similar point estimate for GxE interaction in the VEGF core-genes PS-PRS. We do not find significant evidence of effect-measure modification between OSA and the KEGG HIF1 PS-PRS, TNF PS-PRSs, the NFκB PS-PRSs, nor the full CAD PRS. However, we find a nominally significant trend for risk-amplifying effect-measure modification for both the HIF1 core-genes PS-PRS (interaction OR 1.1, *p* = 4.46E-02) and the curated 225 gene PS-PRS based on a literature-derived CAD- and OSA-involved genes (interaction OR 1.1, *p* = 1.8E-02).

Table 3 reports secondary analysis of module and gene-specific interactions. Adjusting for multiple testing, we find statistically significant effect-measure modification in one of the 30 pathway submodules (VEGF endothelial migration: *CDC42, MAPK11, MAPK12, MAPK13, MAPK14, MAPKAPK2, MAPKAPK3, PTK2, PXN*) and none of the 268 genes tested from the four KEGG pathways of interest. We see nominally significant effect-measure modification in several pathway modules and genes, with estimated effects generally consistent with pathways and modules that contain them (Table 3).

**Table 3:** Secondary Analysis: Genes and Submodules of selected CAD pathways: Covariate-adjusted odds of CAD per s.d. of standardized PS-PRSs, with effect modification by OSA status. Table 3. Analysis of module and gene-level estimates of genetic effects in OSA cases and controls, presented as a secondary post-hoc analysis suggestively localizing primary analysis results. In section A, each core-gene module, defined as the nominal gene and its receptors, is accompanied by each of its constituent genes; in section B, each KEGG pathway is accompanied with those submodules that showed nominally significant main and interaction effects; in section C, genes with the largest risk-inverting and risk-amplifying estimated interaction effects, having nominally significant main effects, are presented. Core-gene modules and KEGG pathways were allocated type I error in the primary analysis. Secondary analysis is underpowered to comprehensively identify driver genes and modules. Nominally significant GxE results (*) do not pass Bonferroni (**) thresholds for multiple testing for 30 pathway modules (1.67E-3), and 268 genes (1.87E-4) with membership in the KEGG HIF1, VEGF, NFκB or TNF pathways. Comprehensive secondary analysis gene- and module-level results for these KEGG pathways are presented in Supplementary Table S4. OR: odds ratio; CI: confidence interval; OSA: Obstructive sleep apnea, CAD: coronary artery disease, KEGG: Kyoto encyclopedia of genes and genomes, HIF1: hypoxia inducible factor 1, VEGF: vascular endothelial growth factor, NFκB: nuclear factor kappa-beta, TNF: tumor necrosis factor, PRS: polygenic risk score, PS-PRS: pathway-specific polygenic risk score.

| A) Core-gene modules with all constituent genes |  |  |  |  |  |  |  |  |  |  |
| --- | --- | --- | --- | --- | --- | --- | --- | --- | --- | --- |
|  | OSA |  | PS-PRS in OSA Controls |  |  | PS-PRS in OSA Cases |  |  | OSA x PS-PRS Interaction |  |
|  | OR | CI | OR | CI | p-value | OR | CI | p-value | OR | p-value |
| ***HIF1 core genes*** | **1.50** | **(1.38, 1.64)** | **1.022** | **(1.009, 1.035)** | **1.06E-03** | **1.114** | **(1.025, 1.212)** | **1.10E-02** | **1.09** | **4.46E-02\*** |
| *HIF1A* | 1.51 | (1.38, 1.64) | 1.005 | (0.992, 1.018) | 4.59E-01 | 0.978 | (0.899, 1.063) | 5.95E-01 | 0.973 | 5.21E-01 |
| *ARNT* | 1.50 | (1.38, 1.64) | 1.022 | (1.009, 1.035) | 1.10E-03 | 1.115 | (1.025, 1.212) | 1.08E-02 | 1.091 | 4.36E-02* |
| ***VEGF core genes*** | **1.51** | **(1.38, 1.64)** | **1.032** | **(1.019, 1.045)** | **1.66E-06** | **0.952** | **(0.874, 1.037)** | **2.58E-01** | **0.92** | **6.70E-02** |
| *VEGFA* | 1.51 | (1.38, 1.64) | 1.018 | (1.005, 1.031) | 7.36E-03 | 0.941 | (0.866, 1.022) | 1.50E-01 | 0.924 | 6.52E-02 |
| *FLT1* | 1.51 | (1.38, 1.64) | 1.030 | (1.017, 1.043) | 7.73E-06 | 0.961 | (0.882, 1.046) | 3.56E-01 | 0.933 | 1.14E-01 |
| *KDR* | 1.51 | (1.38, 1.64) | 1.002 | (0.989, 1.015) | 7.56E-01 | 0.972 | (0.894, 1.057) | 5.09E-01 | 0.970 | 4.83E-01 |
| ***NFκB core genes*** | **1.51** | **(1.38, 1.64)** | **1.031** | **(1.018, 1.045)** | **4.45E-06** | **0.981** | **(0.903, 1.066)** | **6.54E-01** | **0.95** | **2.45E-01** |
| *NFKBIA* | 1.51 | (1.38, 1.64) | 1.005 | (0.992, 1.018) | 4.69E-01 | 1.010 | (0.927, 1.100) | 8.28E-01 | 1.005 | 9.16E-01 |
| *NFKB1* | 1.51 | (1.38, 1.64) | 1.006 | (0.993, 1.019) | 4.05E-01 | 0.938 | (0.862, 1.019) | 1.30E-01 | 0.932 | 1.05E-01 |
| *RELA* | 1.51 | (1.38, 1.64) | 1.025 | (1.012, 1.039) | 2.20E-04 | 1.003 | (0.923, 1.089) | 9.51E-01 | 0.978 | 6.04E-01 |
| *NFKB2* | 1.51 | (1.38, 1.64) | 1.018 | (1.004, 1.031) | 9.02E-03 | 1.076 | (0.990, 1.170) | 8.51E-02 | 1.058 | 1.95E-01 |
| *RELB* | 1.50 | (1.38, 1.64) | 1.015 | (1.001, 1.028) | 2.89E-02 | 0.914 | (0.842, 0.992) | 3.16E-02 | 0.901 | 1.37E-02* |
| ***TNF core genes*** | **1.51** | **(1.38, 1.64)** | **1.019** | **(1.006, 1.033)** | **4.44E-03** | **1.013** | **(0.931, 1.102)** | **7.59E-01** | **0.99** | **8.94E-01** |
| *TNF* | 1.51 | (1.38, 1.64) | 1.013 | (1.000, 1.027) | 4.59E-02 | 1.022 | (0.941, 1.110) | 6.07E-01 | 1.009 | 8.43E-01 |
| *TNFRSF1A* | 1.51 | (1.38, 1.64) | 1.014 | (1.000, 1.027) | 4.32E-02 | 1.008 | (0.927, 1.097) | 8.46E-01 | 0.995 | 9.05E-01 |
| *TNFRSF1B* | 1.51 | (1.38, 1.64) | 1.014 | (1.001, 1.027) | 3.53E-02 | 1.010 | (0.928, 1.098) | 8.24E-01 | 0.996 | 9.18E-01 |

| B) KEGG pathway sub-modules with suggestive GxE |  |  |  |  |  |  |  |  |  |  |
| --- | --- | --- | --- | --- | --- | --- | --- | --- | --- | --- |
|  | OSA |  | PS-PRS in OSA Controls |  |  | PS-PRS in OSA Cases |  |  | OSA x PS-PRS Interaction |  |
|  | OR | CI | OR | CI | p-value | OR | CI | p-value | OR | p-value |
| HIF1 module:<br>IH-induced<br>HIF1-α<br>degradation | 1.51 | (1.38, 1.64) | 1.032 | (1.019, 1.045) | 1.82E-06 | 0.928 | (0.853, 1.009) | 8.05E-02 | 0.899 | 1.42E-02* |
| HIF1 module:<br>angiogenesis | 1.51 | (1.38, 1.64) | 1.032 | (1.019, 1.046) | 1.21E-06 | 0.947 | (0.869, 1.032) | 2.18E-01 | 0.918 | 5.25E-02 |
| VEGF module:<br>endothelial<br>migration | 1.51 | (1.39, 1.65) | 1.014 | (1.001, 1.027) | 3.66E-02 | 0.881 | (0.810, 0.958) | 3.05E-03 | 0.869 | 1.16E-03** |
| VEGF module:<br>endothelial<br>permeability | 1.51 | (1.38, 1.64) | 1.020 | (1.007, 1.034) | 3.20E-03 | 0.913 | (0.838, 0.994) | 3.63E-02 | 0.895 | 1.17E-02* |
| VEGF module:<br>endothelial<br>survival | 1.51 | (1.38, 1.64) | 1.013 | (1.000, 1.027) | 4.47E-02 | 0.911 | (0.837, 0.990) | 2.88E-02 | 0.899 | 1.35E-02* |
| HIF1&VEGF:<br>shared<br>PI3K/AKT<br>module | 1.51 | (1.38, 1.64) | 1.019 | (1.006, 1.033) | 4.79E-03 | 0.912 | (0.837, 0.992) | 3.23E-02 | 0.894 | 1.08E-02* |

| C) Top five genes from KEGG HIF1, VEGF, NFκB or TNF pathways suggestive of risk-inverting GxE |  |  |  |  |  |  |  |  |  |  |
| --- | --- | --- | --- | --- | --- | --- | --- | --- | --- | --- |
|  | OSA |  | PS-PRS in OSA Controls |  |  | PS-PRS in OSA Cases |  |  | OSA x PS-PRS Interaction |  |
|  | OR | CI | OR | CI | p-value | OR | CI | p-value | OR | p-value |
| *RELB* | 1.50 | (1.38, 1.64) | 1.015 | (1.001, 1.028) | 2.89E-02 | 0.914 | (0.842, 0.992) | 3.16E-02 | 0.901 | 1.37E-02* |
| *PIK3R3* | 1.50 | (1.38, 1.64) | 1.02 | (1.007, 1.033) | 2.51E-03 | 0.928 | (0.843, 1.021) | 1.27E-01 | 0.910 | 5.63E-02 |
| *IRF1* | 1.51 | (1.38, 1.64) | 1.026 | (1.013, 1.040) | 9.50E-05 | 0.944 | (0.869, 1.025) | 1.71E-01 | 0.919 | 4.99E-02 |
| *VEGFA* | 1.51 | (1.38, 1.64) | 1.018 | (1.005, 1.031) | 7.36E-03 | 0.941 | (0.866, 1.022) | 1.50E-01 | 0.924 | 6.52E-02 |
| *PLCG1* | 1.51 | (1.38, 1.64) | 1.023 | (1.010, 1.037) | 4.25E-04 | 0.954 | (0.877, 1.038) | 2.71E-01 | 0.932 | 1.06E-01 |
| D) Top five genes from KEGG HIF1, VEGF, NFκB or TNF pathways suggestive of risk-amplifying GxE |  |  |  |  |  |  |  |  |  |  |
|  | OSA |  | PS-PRS in OSA Controls |  |  | PS-PRS in OSA Cases |  |  | OSA x PS-PRS Interaction |  |
|  | OR | CI | OR | CI | p-value | OR | CI | p-value | OR | p-value |
| *ARNT* | 1.50 | (1.38, 1.64) | 1.022 | (1.009, 1.035) | 1.10E-03 | 1.115 | (1.025, 1.212) | 1.08E-02 | 1.091 | 4.36E-02* |
| *MAPK1* | 1.50 | (1.38, 1.64) | 1.015 | (1.002, 1.028) | 2.70E-02 | 1.095 | (1.007, 1.191) | 3.44E-02 | 1.079 | 7.92E-02 |
| *PDHB* | 1.50 | (1.38, 1.64) | 1.015 | (1.002, 1.028) | 2.69E-02 | 1.088 | (1.000, 1.183) | 5.05E-02 | 1.072 | 1.11E-01 |
| *NFKB2* | 1.51 | (1.38, 1.64) | 1.018 | (1.004, 1.031) | 9.02E-03 | 1.076 | (0.990, 1.170) | 8.51E-02 | 1.058 | 1.95E-01 |
| *IL6R* | 1.51 | (1.38, 1.64) | 1.033 | (1.020, 1.046) | 9.71E-07 | 1.091 | (1.002, 1.187) | 4.45E-02 | 1.056 | 2.13E-01 |

### Sensitivity Analyses

We obtained similar results in sensitivity analyses. For the analyses of males-only, self-reported whites-only, and when adjusting for potentially mediating comorbidities, the KEGG VEGF PS-PGRS had consistently Bonferroni-significant risk-inverting effect-measure modification, while the curated CAD- and OSA-related pathway had consistently nominally significant risk-amplifying effect modification; females-only was underpowered with only 71 joint OSA/CAD cases, nonetheless KEGG VEGF was nominally significant with risk-inverting effect.

## DISCUSSION

### Overview and context

Using biobank-scale data we tested the hypothesis that genetic variation in specific molecular pathways that modulate risk of CAD may influence the propensity for CAD differently in individuals with and without OSA—an exposure postulated to have complex effects on atherosclerosis, inflammation, and angiogenesis. We demonstrate novel interaction effects between OSA and PS-PRS within one pathway (VEGF) postulated *a priori* from current biological understanding to play a role in OSA-related hypoxia in CAD. The effect-measure modification of PS-PRS effects suggests that OSA can increase or attenuate genetic risk for CAD in a pathway-dependent manner, and, reciprocally, that subgroups with specific genetic profiles within certain CAD pathways may experience the influence of OSA differently. Our results also suggest heterogeneity of this effect modification across pathways, modules and genes.

Prior work has investigated phenotypic factors, biomarkers, and genetic variation to understand better and dissect the heterogenous effects of OSA on CAD, including multiple variables, such as age, gender and comorbid diagnoses that may moderate associations of OSA and CVD, as well as blood-based biomarkers of CVD [45, 46]. Nonetheless, prior research using genetic variation to explore the divergent individual responses to hypoxic conditions has been limited, despite suggestions advocating this approach to risk stratification of patients with OSA [47]. Our use of PS-PRS, applied to core gene modules and pathways, provides a novel means for interrogating pathway-level effect-measure modification in large-scale data to uncover biological heterogeneity.

### Primary and secondary analyses

In our primary analysis, we see Bonferroni-significant evidence of risk-inverting effect-measure modification in the KEGG VEGF pathway. In secondary analysis also see suggestive evidence of risk-inverting effect-measure modification across multiple independent signals within this pathway. This qualitative effect-measure modification in the VEGF KEGG pathway suggests that among OSA cases, the direction of effect of the VEGF PS-PRS risk alleles, averaged across markers in the pathway, is reversed compared with OSA controls. For example, among OSA cases whose VEGF risk score is one standard deviation above the mean, we estimate that odds of CAD is decreased by approximately 14% compared to those with the mean VEGF risk score.

One explanation consistent with these data would be that OSA perturbs the tissue environment such that SNVs that increase risk in normoxia on average now function to reduce risk under hypoxia. For example, downregulation of a certain gene under normoxia may have been deleterious, but becomes beneficial under hypoxia (or vice versa). The 39 genes in the KEGG VEGF pathway mediate inflammation and angiogenesis, with genes regulating endothelial cell survival, proliferation, and migration as well as vascular permeability. Under intermittent hypoxia, angiogenesis may predominate, potentially conferring protection from clinically observed or fatal CAD events (ascertained in our CAD outcome) due to heart muscle collateralization.

The KEGG VEGF risk-inverting GxE effects appear to be largely driven by the VEGF endothelial migration submodule. However, there appear to be two additional congruent independent signals contributing the overall KEGG VEGF pathway effect: the VEGF core-genes risk-inverting effects, and a separate independent signal distributed among the VEGF endothelial permeability, endothelial survival and PI3K/AKT submodules, which are correlated to one another due to shared genes. This suggests the utility of pathway level analyses in circumstances where consistent effects are combined across many statistically independent genes or loci, which do not reach significance thresholds on their own, but contribute signal to detection of GxE at the pathway level.

By contrast we see nominally-significant suggestive evidence of risk-amplifying effect-modification in the HIF1 core genes module, apparently driven by the *ARNT* gene, as well as in the PS-PRS consisting of 225 curated OSA-CAD genes, suggesting a risk-amplifying GxE signal may be more widely distributed. If confirmed these effects are consistent with the idea that OSA-related physiological stressors may amplify the deleterious effects certain genetic variants on CAD. For example, *ARNT,* whose gene-product HIF1-*β* creates a heterodimer with HIF-1*α* enabling nuclear activation of the HIF1 transcription factor, is a plausible key player in the regulatory network immediately upstream of a host of cellular responses to hypoxia. Although *ARNT* is not under direct hypoxic regulation, concentration of HIF1-*β* may serve as a rate-limiter on HIF1’s transcription factor activity specifically under hypoxic conditions when HIF1-*α* concentration is maximized [48]. Hence, genetic effects on *ARNT* plausibly interact mechanistically with IH-induced effects on HIF1-*α*. Further investigation of OSA’s and IH’s potential for risk-amplifying effect modification appears warranted. If confirmed, differential positive and negative GxE interaction effects, increasing or decreasing individuals’ prior genetic risk in separate genes and pathways in OSA, would be consistent with the idea of a “double-edged” role of OSA in CAD.

Note that the KEGG HIF-1 pathway contains sub-modules with contrasting GxE effects including the KEGG HIF1 angiogenesis (risk-inverting) and the HIF1 core-genes (risk-amplifying) modules. As observed Supplementary Figure S3, the risk scores for these gene modules are both positively correlated with the KEGG HIF1 PS-PRS as a whole, being component terms in the sum. This observation suggests that the KEGG HIF1 signaling pathway may contain opposing signals which together attenuate overall HIF1 pathway effect modification. This situation illustrates the limitations of targeting large heterogeneous pathways in studying GxE effect modification and underscores the utility of also examining pathways limited to genes or sub-modules that better ensure specificity. Nonetheless, since pathway boundaries can be vague and subjective, further research is needed to investigate data-driven methods for aggregating co-regulated genes to modules and pathways that may respond similarly to stimuli such as IH.

A natural question is whether the strong biological relationships among the chosen pathways challenge our interpretation. Clearly the selected pathways implicated in both IH and CAD are co-regulated: notably VEGF is explicitly regulated by HIF1, but cross-talk and co-regulation among all four pathways including NFκB and TNF likely exists, and gene expression levels may be correlated as a result. Nevertheless, our analysis does not directly analyze gene expression but rather genetic variation and its perturbation of this signaling network due to risk alleles specific to each pathway that are largely statistically independent across pathways. Even though increased concentrations of HIF1 in OSA is expected to augment VEGF, a number of considerations could explain why the CAD effects of e.g. of *ARNT* risk alleles may (on average) be exacerbated but the effects of VEGF pathway risk alleles may (on average) be reversed, including differential downstream effects.

These results open the way to varied experimental studies to probe the potentially novel molecular mechanisms that may lead to the observed qualitative effect-measure modification and underly the findings. Further study of genes and variants in this pathway and their effect on measured biomarkers and gene expression may provide clarification of the genetic mediators of CAD risk in OSA, and potentially point to molecular targets for ameliorating OSA-related CAD. ***Additional Findings***

Testing our primary hypothesis yielded additional interesting findings. We demonstrated that a genome-wide CAD PRS associated with CAD in both the OSA cases and controls was distributed similarly (in mean and standard deviation) and did not reveal significant effect-measure modification of the genome-wide CAD PRS. We also confirmed that the chosen PS-PRSs for CAD associate positively with CAD in OSA controls, and that we see similar distribution of PRS and PS-PRSs when stratifying by OSA status. Additionally, given that nominally significant point estimates of interaction between genetic risk for CAD and OSA differed qualitatively across pathways, this suggests analyses ignoring heterogeneity (such as only assessing the full PRS) may incorrectly lead to the conclusion that there is no evidence of GxE interaction. Lastly, we demonstrated a consistent positive association between OSA and CAD, after adjusting for covariates as well as PS-PRS main and GxE interaction effects, with an effect size (OR 1.5) similar to that previously reported in observational cohort studies [49].

### Strengths and weaknesses

This study has several notable strengths. Our methodology offers the opportunity to detect distinct positive and negative interaction effects in separate pathways, which might otherwise interfere with each other and prevent detection of a GxE interaction at the genome-wide PRS level. Furthermore, if our hypothesis holds, environments such as OSA-related hypoxemia will perturb gene activity across regulatory pathways or submodules containing thousands of SNVs. So, we expect aggregating SNV risk to genes, modules, or pathways acting together to produce similar downstream biological effects may enhance power and reduce the multiple testing burden, as compared with SNV-level interaction testing. So, PS-PRS may provide useful intermediate levels between individual genetic markers and genome-wide risk scores to study effect-measure modification. And unlike the challenges to studying these regulatory pathways with biological assays in relevant tissues and with appropriate timing, our approach extracts additional insights from existing GWAS and biobank data.

The study also has several limitations. First, we perform a cross-sectional analysis, due to the limited follow-up for incident CAD, as well as the difficulty determining the date of onset of sleep apnea. Cross-sectional analysis is commonly used in genetics, including the GWAS and PRS analyses on which our PS-PRS were based. While in the GxE context ideally the OSA exposure should be established as preceding the CAD outcome, the presence of OSA in the medical record generally is believed to reflect disease that occurred years earlier [50]. Second, UK Biobank has several sources of selection bias, including healthy volunteer bias, which may limit generalizability. Also, to overcome a limitation of the UK Biobank dataset in which sleep apnea is not subclassified as central or obstructive either in self-reported or medical records data, we attempted to reduce potential CSA cases, which are likely causally downstream of atherosclerosis and CAD, by requiring the presence of self-reported snoring. Third, the number of ascertained OSA cases is low compared to expected population prevalence estimates.

Nonetheless, we expect our ascertainment of OSA to be specific (likely identifying more severe and hypoxic individuals) if not sensitive, and we took additional steps to reduce misclassification of cases and controls using cardinal symptoms of OSA. Also, the use of a binary classification for OSA also did not allow us to characterize OSA according to severity of hypoxemia or sleep fragmentation, which precluded our ability to test dose-response associations or associations with specific OSA stressors.

### Summary

The existence of large-scale genetic data such as the UK Biobank opens up new avenues in understanding genotype-phenotype relationships in the context of different environments, host factors and comorbid diseases. Genome-wide data on genetic risk variants and novel genomic risk scores enabled by new LD-aware PRS algorithms such as LDPred present opportunities to develop robust pathway-level characterization of genetic risk. Building on these novel PS-PRSs we deploy an interaction analysis that allows us to probe relationships between OSA, CAD genetics and CAD outcomes that localize to particular molecular pathways. In turn we can gain a deeper understanding of genetic disease architecture, including modification of pre-existing genetic risk by exposures such as OSA.

In conclusion, this study provides insight into the influence of OSA on CAD and identifies a novel mechanism that may explain the evident heterogeneity of CAD risk among individuals with OSA. Specifically, we find that OSA status can modify pre-existing genetic risk of CAD within the VEGF pathway, and more generally we provide a plausible approach to assess stratified genetic risk for CAD in OSA by illustrating a novel method for understanding pathway-specific genetic risk and its role in GxE interaction. Future larger PS-PGRS GxE studies may allow probing of dose-dependent effects of OSA-related intermittent hypoxia on a comprehensive set of CAD pathways, while further study of these pathways and genes with additional –omics data or animal studies may further clarify our current findings. Having identified genetic pathways whose effect is modified by OSA status, we may also identify individuals with OSA who are most susceptible (or resilient) to CAD, disentangling their genetic risk, their OSA exposure risk, and the interaction between the two. Together these investigations may lead to personalized management of OSA and the ability to target treatment toward specific impacted pathways in those at heightened risk.

## Data Availability

Data available on request from UK Biobank.

## Acknowledgments

This research has been conducted using the UK Biobank Resource (application 6818). We would like to thank the participants and researchers from the UK Biobank who contributed or collected data.

## Sources of Funding

National Heart, Lung, and Blood Institute, NIH; The University of Manchester (Research Infrastructure Fund).

## Disclosures

Dr. Goodman reports grants from National Heart, Lung, and Blood Institute, NIH, during the conduct of the study; Dr. Cade has nothing to disclose; Dr. Shah has nothing to disclose; Dr. Huang has nothing to disclose; Dr. Dashti has nothing to disclose; Dr. Saxena has nothing to disclose; Dr. Rutter reports consulting fees and non-promotional lecture fees from Novo Nordis, outside the submitted work; Dr. Libby was an unpaid consultant to, or involved in clinical trials for Amgen, AstraZeneca, Baim Institute, Beren Therapeutics, Esperion, Therapeutics, Genentech, Kancera, Kowa Pharmaceuticals, Medimmune, Merck, Norvo Nordisk, Merck, Novartis, Pfizer, Sanofi-Regeneron. Dr. Libby is a member of scientific advisory boards for Amgen, Corvidia Therapeutics, DalCor Pharmaceuticals, Kowa Pharmaceuticals, Olatec Therapeutics, Medimmune, Novartis, and XBiotech, Inc during the conduct of this study; Dr. Libby’s laboratory has received research funding in the last two years from Novartis; Dr. Libby also reports grants from the National Heart, Lung, and Blood Institute, the American Heart Association, the RRM Charitable Fund, and the Simard Fund during the conduct of this study; Dr. Libby is on the Board of Directors of XBiotech, Inc. Dr. Libby has a financial interest in Xbiotech, a company developing therapeutic human antibodies; in addition, Dr. Libby has a patent Use of Canakinumab pending to Ridker P, Thuren T, Bermann G, Libby P, inventors; Dr. Libby’s interests were reviewed and are managed by Brigham and Women’s Hospital and Partners HealthCare in accordance with their conflict of interest policies; Dr. Sofer has nothing to disclose. Dr. Redline reports grants from NIH during the conduct of the study; grants and personal fees from Jazz Pharma, personal fees from Eisai Inc, personal fees from Apnimed Inc, outside the submitted work; and Dr. Redline is the first incumbent of an endowed professorship donated to the Harvard Medical School by Dr. Peter Farrell, the founder and Board Chairman of ResMed, through a charitable remainder trust instrument, with annual support equivalent to the endowment payout provided to the Harvard Medical School during Dr. Farrell’s lifetime by the ResMed Company through an irrevocable gift agreement.

## Key Figure

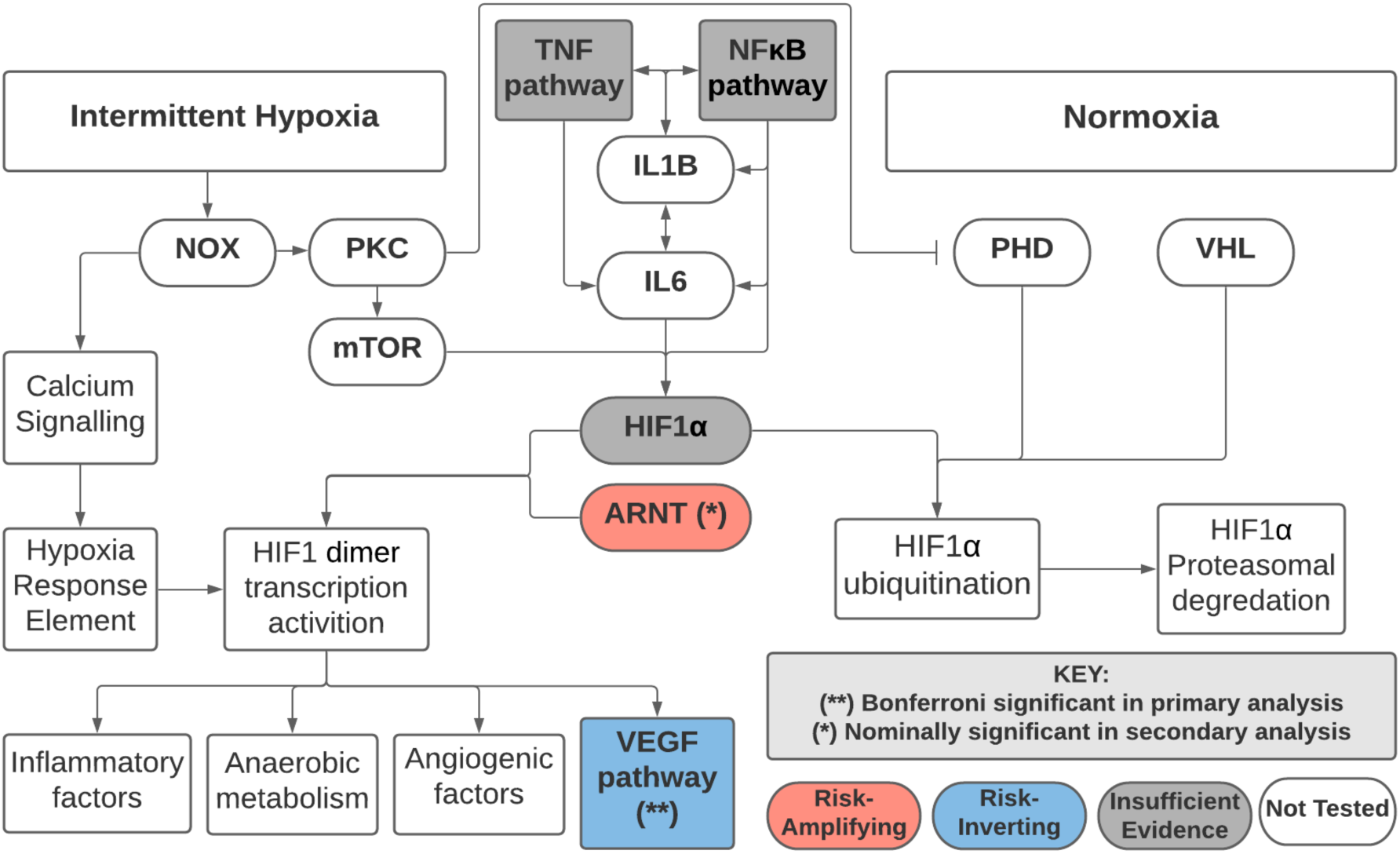

Estimated qualitative effect-measure modification by OSA on genetic risk of CAD, using pathway-specific polygenic risk scores (PS-PRS) and gene-specific risk scores (GSRS), for the given pathways and genes.^2^

## Methods Supplement

### Selecting genomic variants and annotating genes

We obtained 6.6 million common SNVs (minor allele frequency > 0.01) available in UK Biobank imputed data which passed quality control filters in a prior GWAS and subsequent PRS analysis of CAD conducted by Khera et al. These SNVs were then given genetic annotations using a sequential multi-stage methodology as described above in the text and shown in Figure 1.

We ordered the quality of the annotation resources as follows: GenCode v34 exons [39], FANTOM5 [40], Promoter-Capture HiC experiments involving white blood cells as described in [41], GTEx v7 statistically significant eQTLs [42] derived from artery (aorta, coronary and tibial), heart (atrial appendage and left ventricle) and whole blood tissues, GeneHancer v4.4 [43] predicted gene annotations passing minimum enhancer score threshold of 0.3 and minimum gene score threshold of 4.0, and finally GenCode protein-coding regions (including all loci from 20kbp upstream of transcription start site to 10kbp downstream of transcription stop site), and noncoding transcript sequence regions. Within a single data set a SNV may be assigned to multiple genes, in which case both annotations are accepted.

### Calculating genetic risk scores

After defining pathways and associated SNVs, we calculated a PS-PRS for each participant using a SNV-set representing each of the ten pathways (HIF-1, VEGF, NFκB, TNF represented by both a core-gene module and a KEGG pathway, the curated CAD/OSA pathway, and the full PRS). For each PS-PRS, we took the subset of SNVs annotated to that pathway and calculated a PS-PRS using the corresponding genetic effects from the Khera et al PRS. We standardized polygenic risk scores including individual PS-PRSs to mean zero and variance one in the total UKBB study population. To facilitate our secondary analysis, we additionally calculated GSRS for each gene in the HIF-1, VEGF, NFκB, and TNF KEGG pathways.

### Modeling and testing associations

For each PRS we created a single CAD outcome model and tested: A) the association of the PS-PRS with CAD, and B) the interaction of the PS-PRS with OSA status, controlling for demographics and comorbidities. We modeled the binary CAD outcome via generalized additive models (GAM) with logistic link, controlling for the covariates discussed above [44].

Statistical significance was assessed at level *α* = 0.05. For our primary analysis testing interaction of PS-PRS and OSA status, we adopted the Bonferroni-adjusted significance threshold *α* = 0.005 on ten interaction tests. In secondary analysis of GSRS, we report those genes that had risk-amplifying main effects on CAD (in OSA controls) and passed an *α* = 0.05 main effect significance threshold. Statistical calculations were conducted using *R* version 3.5.2, and code used to perform these analyses will be posted to a publicly accessible repository (https://github.com/MatthewOGoodman/UKB_PS-PRSxOSA_GxE_study_of_CAD).

---

**Supplementary Table S1:**
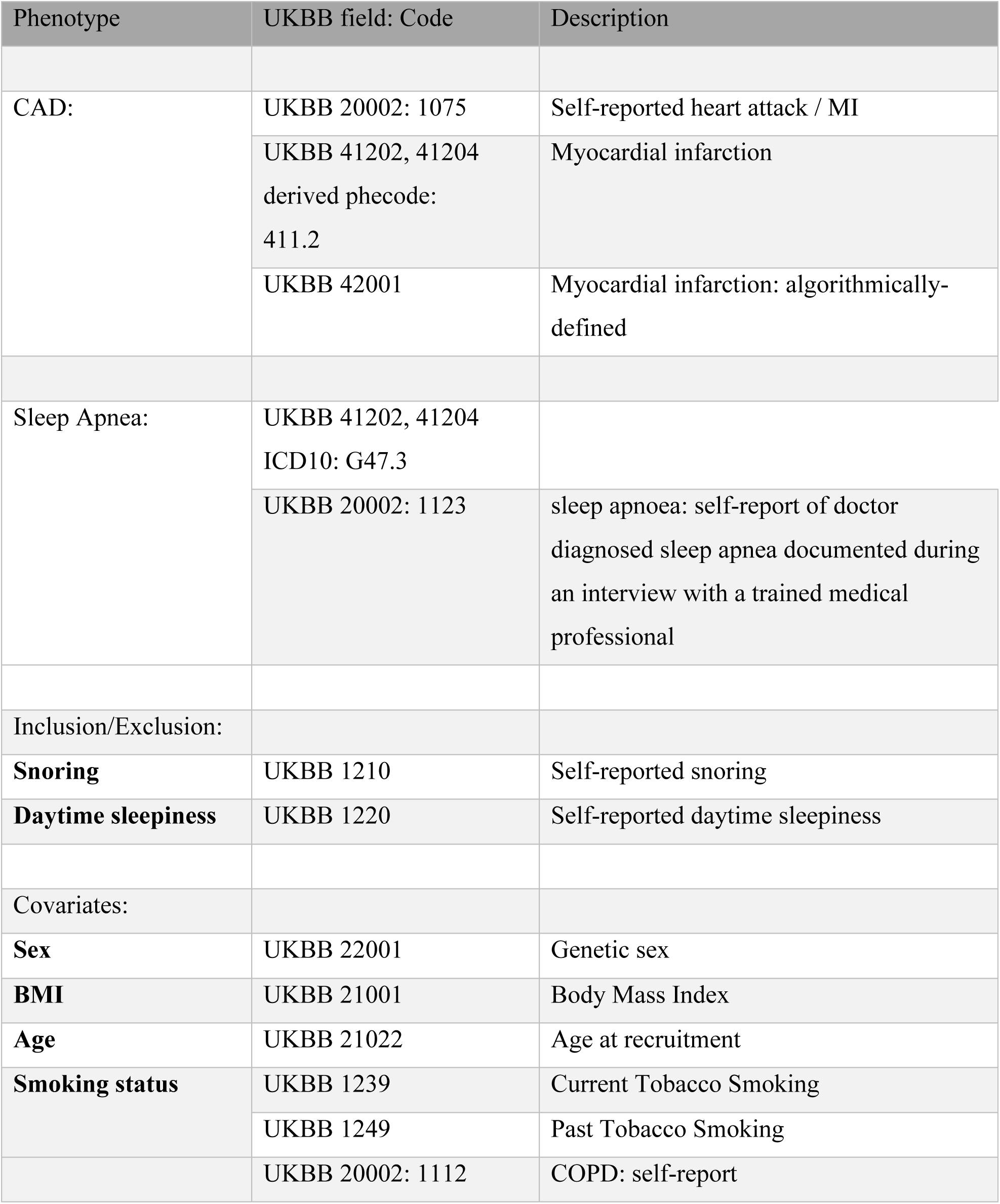
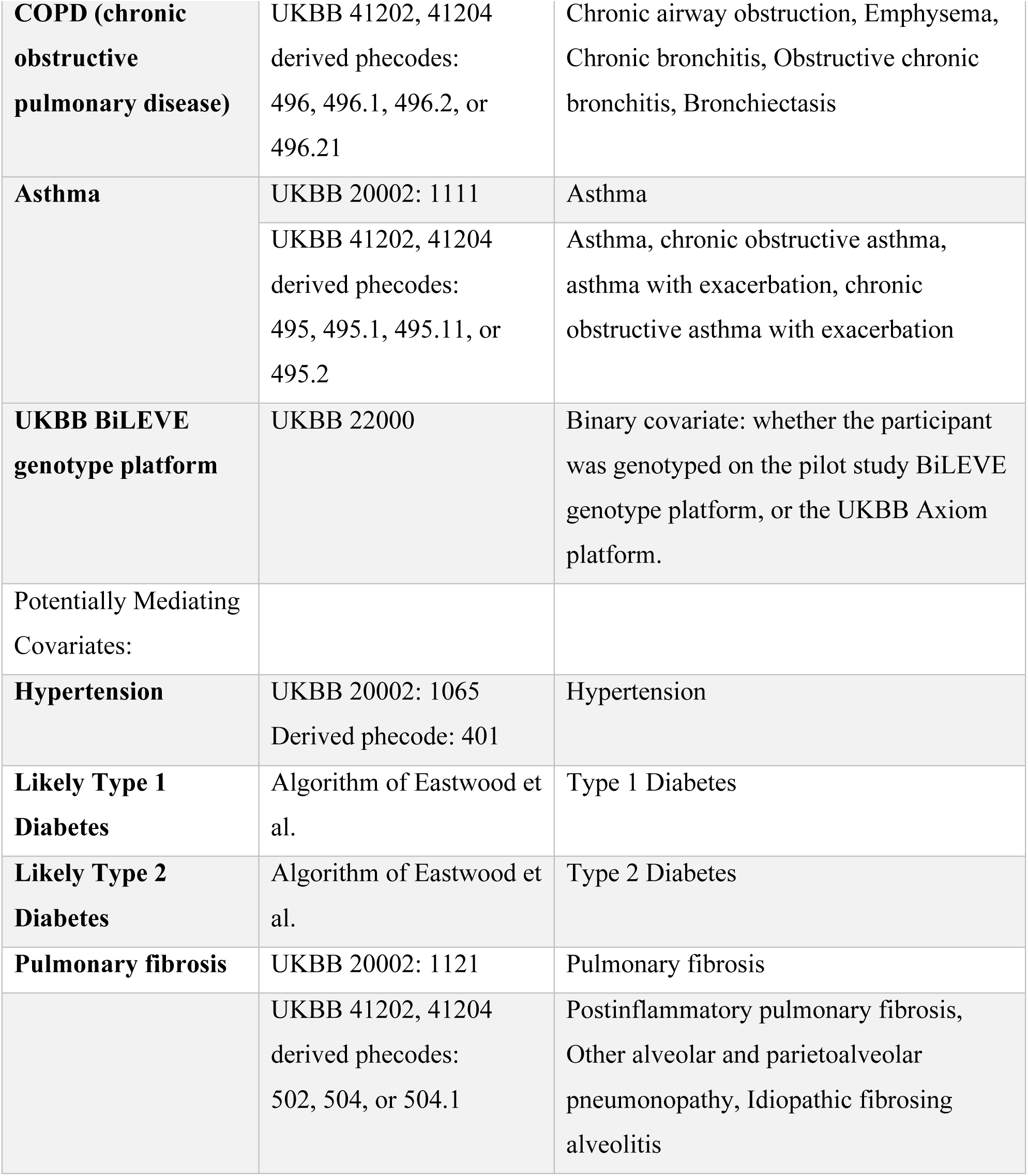
Phenotype definitions and data sources Daytime sleepiness was recoded as “present” (moderate or severe) or “absent” (none or mild). Smoking status was recoded as “never smoked,” “past smoker,” “current smoker (occasionally),”and “current smoker (most days).” Diagnostic information for medical diagnoses was obtained from the following UKBB data fields: Non-cancer illness, self-reported (20002), Hospital Episode Inpatient data: Diagnoses - main ICD10 (41202), Diagnoses - secondary ICD10 (41204), ICD10 codes were collapsed to medically interpretable groupings using the phecode system using Phecode Map version 1.2 for the World Health Organization ICD10 codings.

| Phenotype | UKBB field: Code | Description |
| --- | --- | --- |
| CAD: | UKBB 20002: 1075 | Self-reported heart attack / MI |
|  | UKBB 41202, 41204<br>derived phecode:<br>411.2 | Myocardial infarction |
|  | UKBB 42001 | Myocardial infarction: algorithmically-defined |
| Sleep Apnea: | UKBB 41202, 41204<br>ICD10: G47.3 |  |
|  | UKBB 20002: 1123 | sleep apnoea: self-report of doctor diagnosed sleep apnea documented during an interview with a trained medical professional |
| Inclusion/Exclusion: |  |  |
| **Snoring** | UKBB 1210 | Self-reported snoring |
| **Daytime sleepiness** | UKBB 1220 | Self-reported daytime sleepiness |
| Covariates: |  |  |
| **Sex** | UKBB 22001 | Genetic sex |
| **BMI** | UKBB 21001 | Body Mass Index |
| **Age** | UKBB 21022 | Age at recruitment |
| **Smoking status** | UKBB 1239 | Current Tobacco Smoking |
|  | UKBB 1249 | Past Tobacco Smoking |
|  | UKBB 20002: 1112 | COPD: self-report |
| **COPD (chronic obstructive pulmonary disease)** | UKBB 41202, 41204 derived phecodes: 496, 496.1, 496.2, or 496.21 | Chronic airway obstruction, Emphysema, Chronic bronchitis, Obstructive chronic bronchitis, Bronchiectasis |
| **Asthma** | UKBB 20002: 1111 | Asthma |
|  | UKBB 41202, 41204 derived phecodes: 495, 495.1, 495.11, or 495.2 | Asthma, chronic obstructive asthma, asthma with exacerbation, chronic obstructive asthma with exacerbation |
| **UKBB BiLEVE genotype platform** | UKBB 22000 | Binary covariate: whether the participant was genotyped on the pilot study BiLEVE genotype platform, or the UKBB Axiom platform. |
| Potentially Mediating Covariates: |  |  |
| **Hypertension** | UKBB 20002: 1065<br>Derived phecode: 401 | Hypertension |
| **Likely Type 1 Diabetes** | Algorithm of Eastwood et al. | Type 1 Diabetes |
| **Likely Type 2 Diabetes** | Algorithm of Eastwood et al. | Type 2 Diabetes |
| **Pulmonary fibrosis** | UKBB 20002: 1121 | Pulmonary fibrosis |
|  | UKBB 41202, 41204 derived phecodes: 502, 504, or 504.1 | Postinflammatory pulmonary fibrosis, Other alveolar and parietoalveolar pneumonopathy, Idiopathic fibrosing alveolitis |

**Supplementary Table S2:**
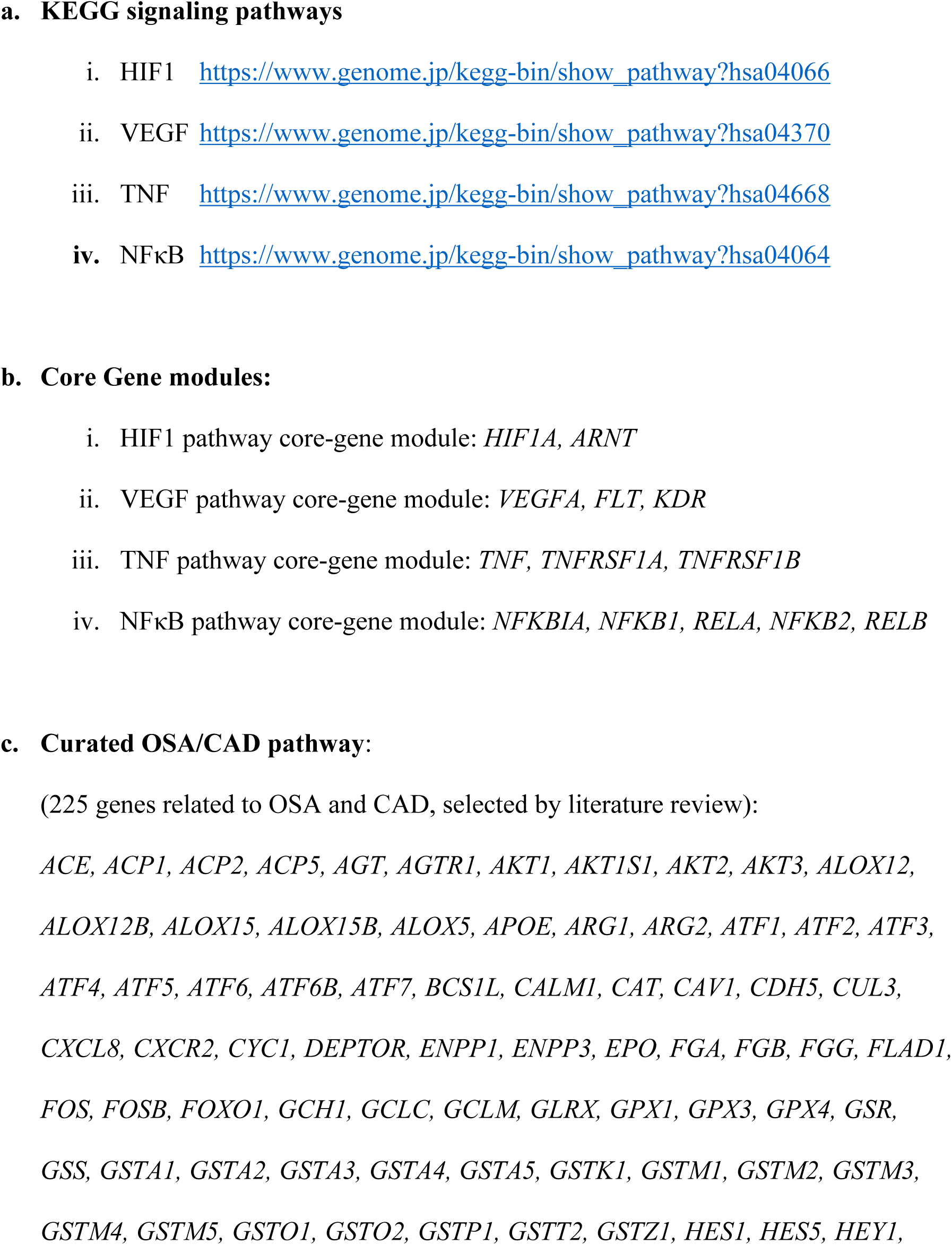
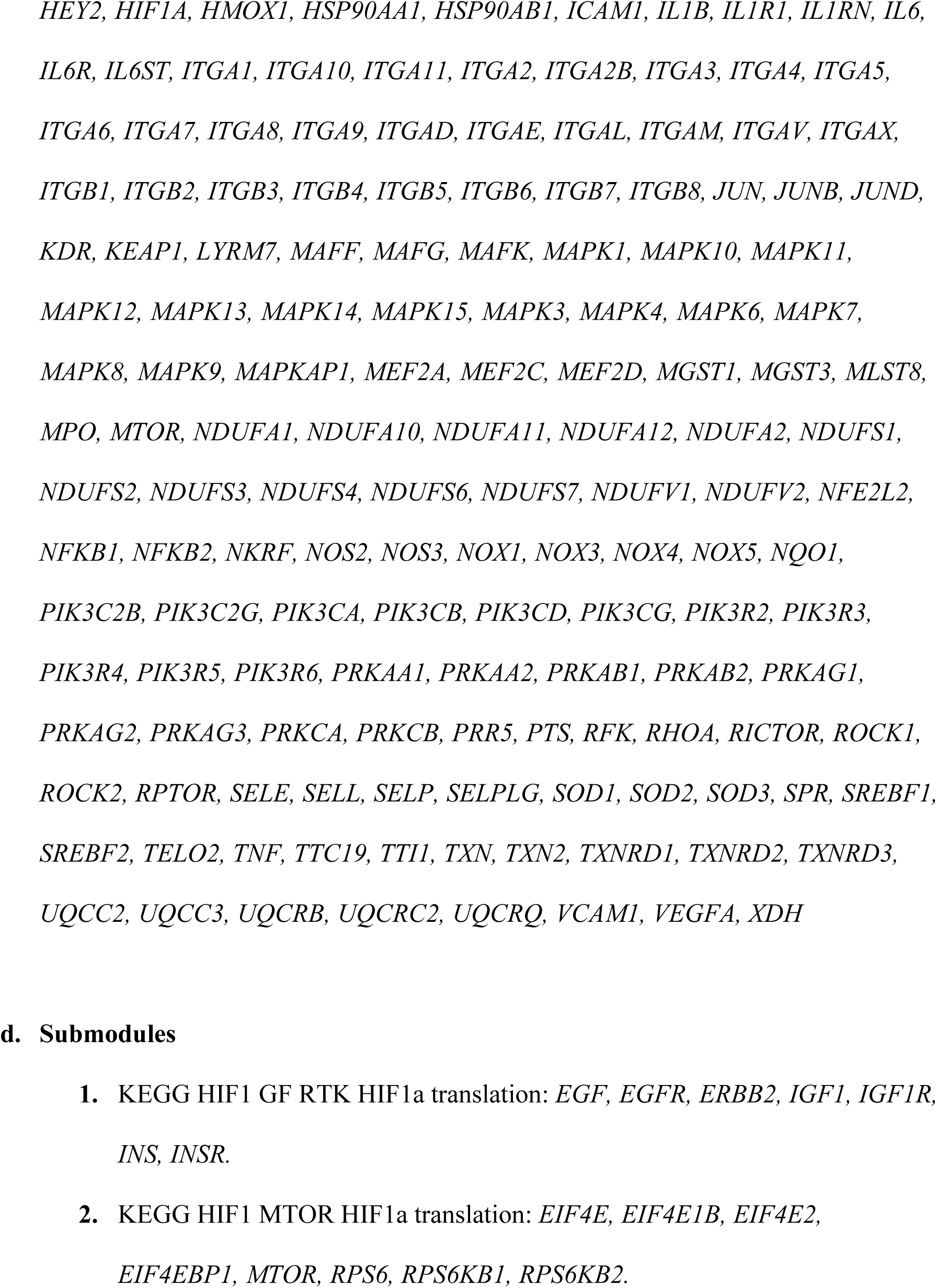
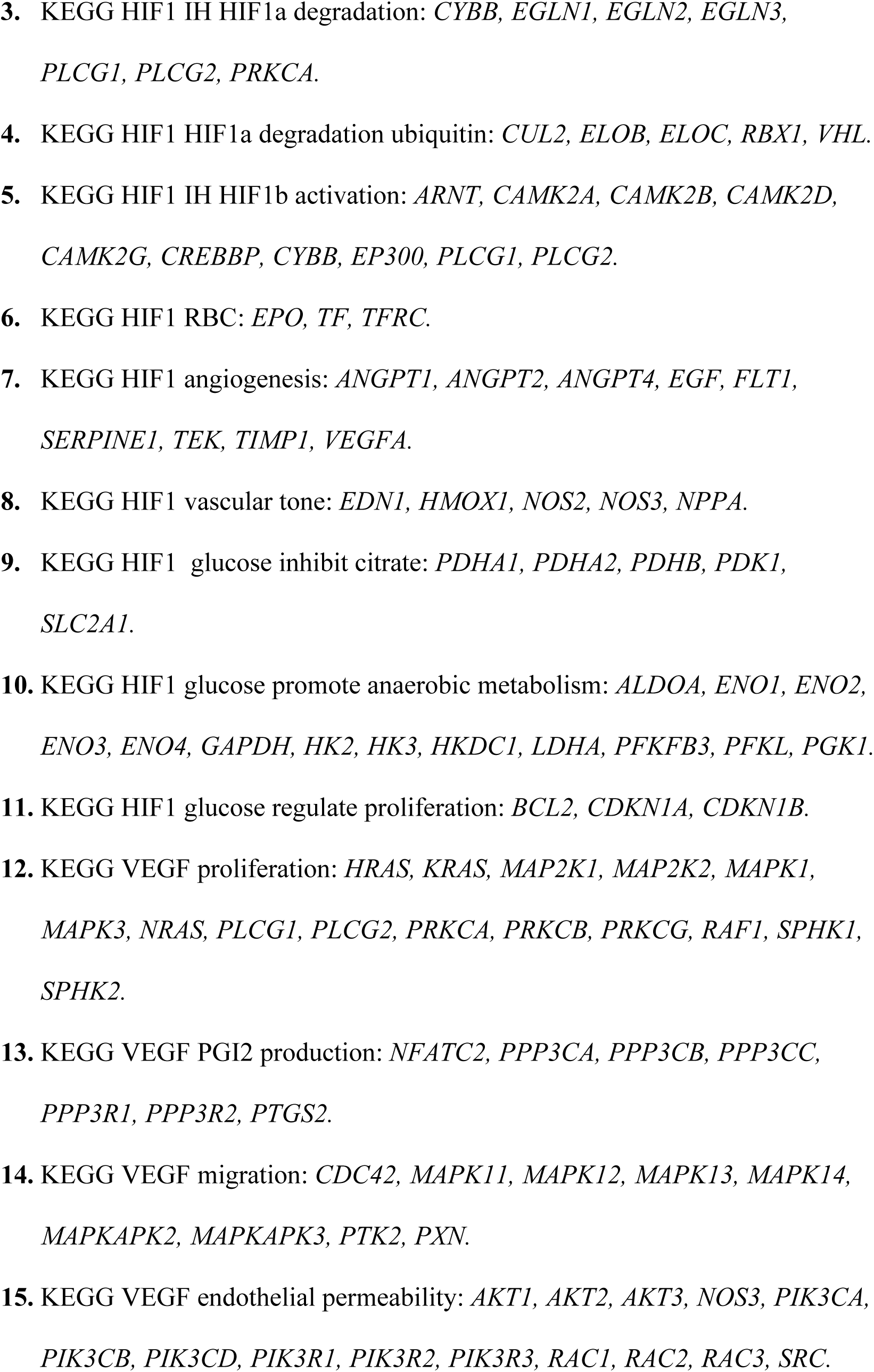
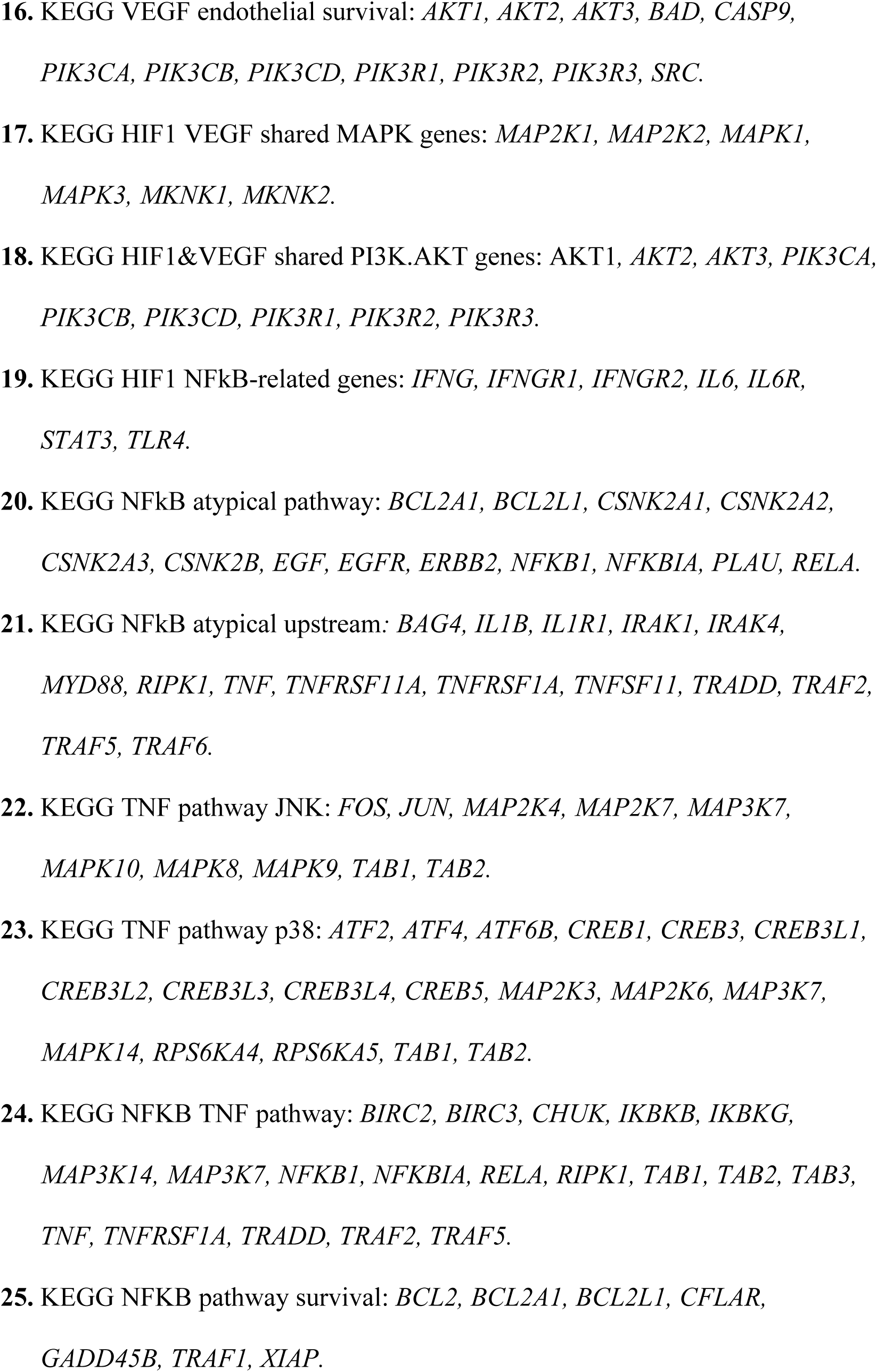
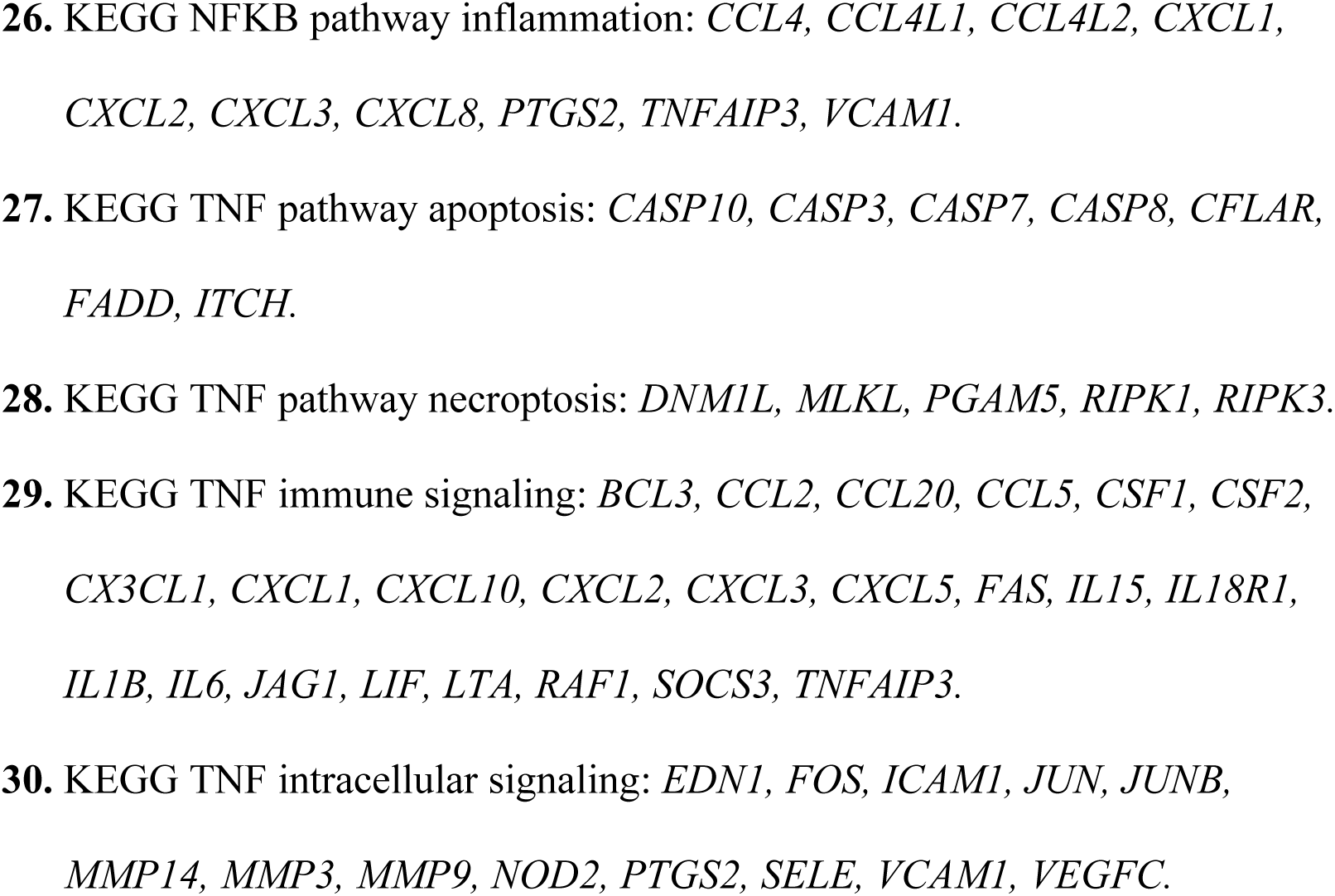
Pathway and module definitions Pathway Gene Membership a. KEGG signaling pathways

**Supplementary Figure S3:**
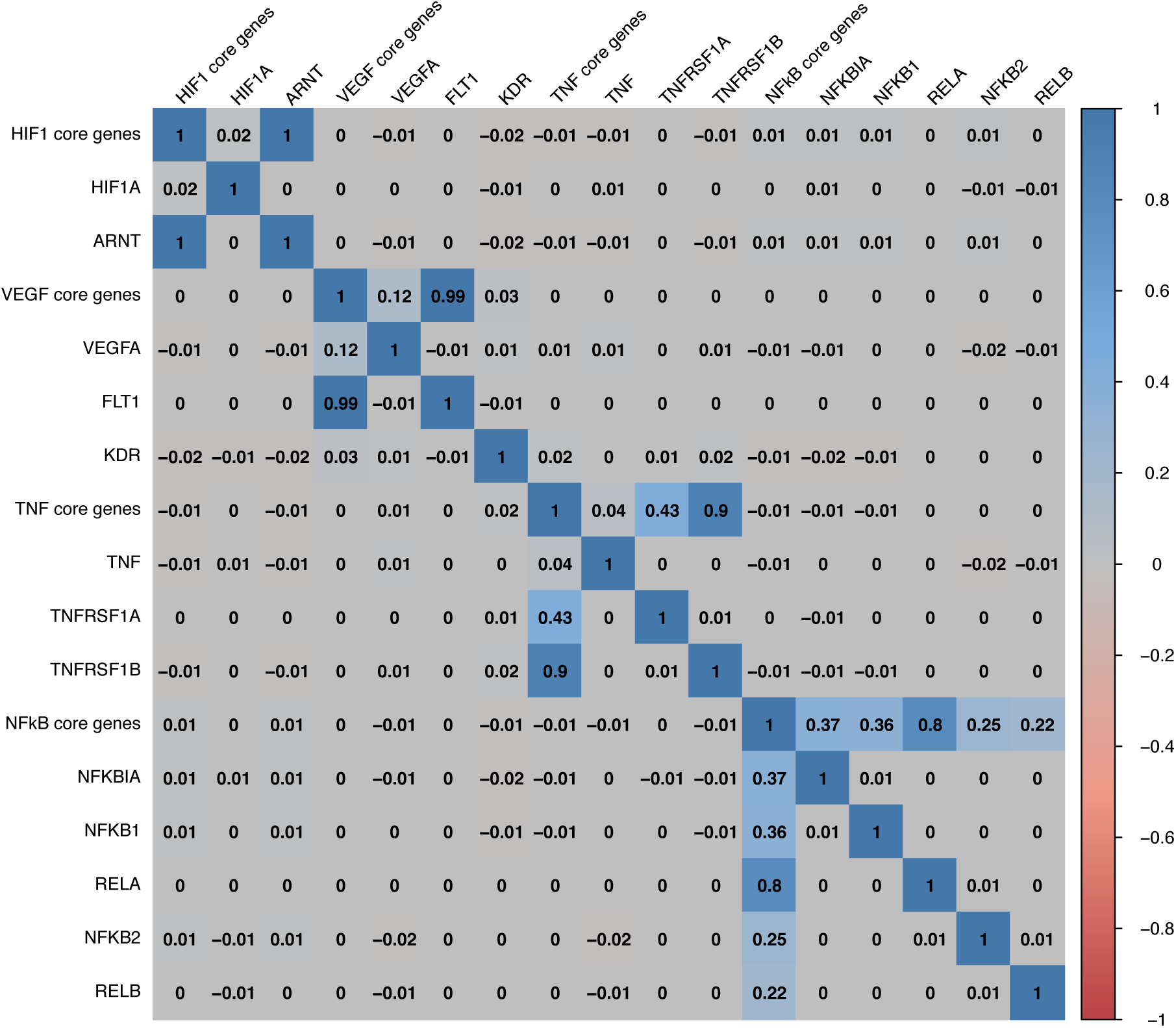
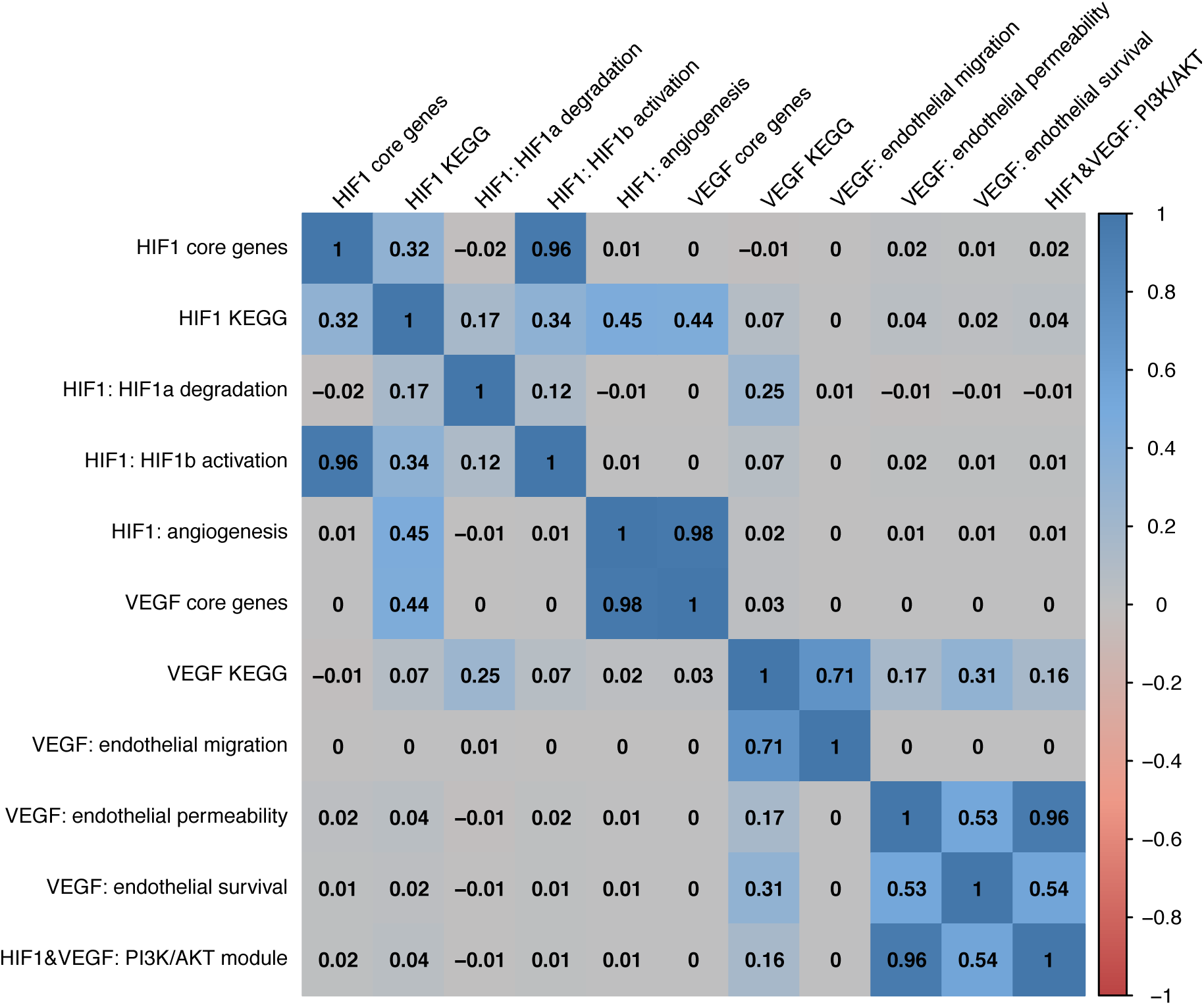
Correlations between pathways, modules and genes. Figure S3a) Pearson correlations between core-gene PS-PRSs and their constituent gene-specific risk scores (GSRS). Components genes are listed below each core-genes pathway. Figure S3b) Pearson correlations between selected HIF1 and VEGF sub-module, KEGG and core-genes PS-PRSs in the secondary analysis. Pathway definitions are shown in Supplementary Table S2.

**Supplementary Table S4:** Effect estimates of covariates for model of full PRS

|  | Est. | s.e. | p-value | OR | OR CI |
| --- | --- | --- | --- | --- | --- |
| (Intercept) | -3.49436 | 0.044507 | 0.00E+00 | 0.0304 | (0.028, 0.033) |
| sex (male) | 1.04334 | 0.045079 | 1.64E-118 | 2.8387 | (2.599, 3.101) |
| smoking (never) | -0.93997 | 0.042274 | 1.56E-109 | 0.3906 | (0.360, 0.424) |
| smoking (occasionally) | -0.31976 | 0.090599 | 4.16E-04 | 0.7263 | (0.608, 0.867) |
| smoking (previous) | -0.63896 | 0.043239 | 2.05E-49 | 0.5278 | (0.485, 0.575) |
| white | 0.06102 | 0.025926 | 1.86E-02 | 1.0629 | (1.010, 1.118) |
| Genetic PC1 (mean (SD)) | 0.00242 | 0.000192 | 2.71E-36 | 1.0024 | (1.002, 1.003) |
| Genetic PC2 (mean (SD)) | -0.00078 | 0.000318 | 1.41E-02 | 0.9992 | (0.999, 1.000) |
| Genetic PC3 (mean (SD)) | 0.0072 | 0.000527 | 1.29E-42 | 1.0072 | (1.006, 1.008) |
| Genetic PC4 (mean (SD)) | 0.00173 | 0.000691 | 1.23E-02 | 1.0017 | (1.000, 1.003) |
| Genetic PC5 (mean (SD)) | 0.00857 | 0.000916 | 8.61E-21 | 1.0086 | (1.007, 1.010) |
| BiLEVE | 0.12948 | 0.01998 | 9.13E-11 | 1.1382 | (1.095, 1.184) |
| COPD | 0.79107 | 0.023188 | 4.38E-255 | 2.2058 | (2.108, 2.308) |
| asthma | 0.08522 | 0.019372 | 1.09E-05 | 1.089 | (1.048, 1.131) |
| OSA | 0.41954 | 0.046994 | 4.36E-19 | 1.5213 | (1.387, 1.668) |
| CAD PRS | 0.52797 | 0.00711 | 0.00E+00 | 1.6955 | (1.672, 1.719) |
| sex by smoking (never) | 0.34135 | 0.049628 | 6.06E-12 | 1.4069 | (1.276, 1.551) |
| sex by smoking<br>(occasionally) | -0.0056 | 0.102995 | 9.57E-01 | 0.9944 | (0.813, 1.217) |
| sex by smoking (previous) | 0.38252 | 0.050397 | 3.20E-14 | 1.4660 | (1.328, 1.618) |
| OSA by CAD PRS | -0.01857 | 0.045087 | 6.80E-01 | 0.9816 | (0.899, 1.072) |

**Supplementary Table S5:** Comprehensive secondary analysis GxE testing results for KEGG HIF1, VEGF, NFκB or TNF pathways. (See file Goodman_Supplementary_Tables.csv) A) module-level GxE results B) gene-level GxE results

**Supplementary Table S6:** Sensitivity Analysis results (See file Goodman_Supplementary_Tables.csv) C) Adjusting for potential mediators (hypertension, T1D, T2D and pulmonary fibrosis) D) Males only E) Females Only F) White Europeans only

## Footnotes

1 ICD9 records were not analyzed as A) ICD9 codes come from a unique subset of 20k Scottish participants with available hospital inpatient episodes from 1980 to 1996, whereas the ICD10 data was collected almost exclusively after 1996 B) the fact that these ICD9s contain no instances of sleep apnea codes 327.23 or 327.2, nor any sleep codes in the 327 range.

2 *HIF1α* and *ARNT* constitute the HIF1 core-genes module and had estimated risk-amplifying effect modification at nominal significance in primary analysis. HIF1 KEGG pathway not shown, but encompasses HIF1 core-genes as well as additional genes and modules shown in the graphic, and its estimated PS-PGRS effect modification did not meet nominal significance in primary analysis. Bonferroni-corrected significance level of 0.005 of based on 10 hypothesis tests in primary analysis and nominal level 0.05.

## Notes

### Funding Statement

Dr. Goodman reports grants from the National Heart, Lung, and Blood Institute, NIH; Dr. Rutter reports funding from The University of Manchester (Research Infrastructure Fund)

### Author Declarations

Investigating pathway-specific genetic risk of coronary artery disease and its relationship with intermittent hypoxia in obstructive sleep apnea IR Approved v1.0 PI: Goodman, Matthew (MG185) Sponsor: NIH-National Institutes of Health Agreement #: 2020A010894

